# Spermidine suppresses glial inflammation and parkinsonian abnormalities in ATP13A2 deficiency

**DOI:** 10.64898/2026.05.23.26353575

**Authors:** Ana Cascalho, Aisha Sati, Hanne Dhondt, Nina Schoonvliet, Natalie Kaempf, Elena Coccia, Alexandra Mamalaki, Maria I. Behrens, Norbert Brüggemann, Markus Glatzel, Veerle Baekelandt, Christine Klein, Jan Eggermont, Patrik Verstreken, Joel Blanchard, Peter Vangheluwe

## Abstract

Pathogenic variants in *ATP13A2*, which encodes an endolysosomal polyamine exporter, cause Kufor-Rakeb syndrome and are associated with early-onset parkinsonism and related neurodegenerative disorders; however, the mechanisms by which ATP13A2 dysfunction drives disease remain incompletely defined. In *Atp13a2* knockout mice, we identified an early, transient reduction in brain polyamines that precedes overt gliosis and behavioural abnormalities. Pharmacological polyamine depletion exacerbates phenotypes, whereas oral supplementation of spermidine, but not spermine, rescues parkinsonian symptoms establishing metabolic polyamine deficiency as a pathogenic driver. Mechanistically, spermidine counteracts microglia lysosomal dysfunction in the brain and exerts mitochondrial antioxidant and anti-inflammatory effects in primary mouse microglia, thereby improving neuronal integrity. In the absence of Atp13a2, microglial spermidine import relies on the related polyamine transporter Atp13a3. Importantly, these findings translate to human systems, whereby spermidine attenuates inflammation in ATP13A2-deficient human differentiated microglia, while postmortem ATP13A2-deficient brain analysis confirms increased microglia reactivity. Spermidine also rescues motor deficits and dopaminergic neuron loss in ATP13A2-deficient Drosophila and other fly parkinsonism models. Together, these findings identify early polyamine dysregulation as a mechanistic contributor to ATP13A2-associated parkinsonism and nominate spermidine supplementation as a potential therapeutic strategy for ATP13A2-driven pathology and possibly a broader range of parkinsonian sub-types.

## Introduction

Parkinsonian neurodegeneration is increasingly recognized as a process involving multiple brain cell types, in which glial inflammatory states can precede and amplify neuronal vulnerability^1,2^. Pathogenic loss-of-function variants in *ATP13A2*/*PARK9* cause Kufor-Rakeb syndrome (KRS; PARK-ATP13A2), a rare and severe juvenile-onset form of parkinsonism, and have also been implicated in early-onset Parkinson’s disease (EOPD) and related neurodegenerative disorders^3–6^. Clinically, KRS and *ATP13A2*-linked EOPD are characterized by motor abnormalities, including bradykinesia, rigidity, and early postural instability^4,7^, reflecting dysfunction of the basal ganglia and substantia nigra, which is recapitulated in animal models^1,8^. Reduced ATP13A2 levels have also been reported in dopaminergic neurons of the substantia nigra in sporadic Parkinson’s disease (PD), suggesting that ATP13A2 dysfunction may be relevant to both monogenic and more common forms of parkinsonism^9^.

ATP13A2 encodes an endolysosomal transporter that exports the polyamines spermidine (Spd) and spermine (Spm) from the late endolysosomal lumen to the cytosol, thereby supporting intracellular polyamine homeostasis ^10,11^. Polyamines are essential small polycations with antioxidant and anti-inflammatory properties that regulate diverse cellular processes, including gene expression, protein folding, ion channel activity, autophagy, mitochondrial function, and lysosomal homeostasis, many of which are disrupted in PD^12,13^. Loss of ATP13A2 function causes lysosomal polyamine accumulation, impairing lysosomal degradative capacity, glucocerebrosidase activity, and lysosomal membrane integrity, whereas the impaired lysosomal polyamine flux also sensitizes cells to mitochondrial oxidative stress^9–11,14–17^. Altered polyamine levels have also been reported in plasma and cerebrospinal fluid from patients with sporadic PD and other neurodegenerative diseases, suggesting that polyamine imbalance may play a broader role in neurodegeneration^18,19^.

Despite these links, the mechanisms of how impaired lysosomal polyamine transport may give rise to parkinsonian phenotypes remain unclear. Germline or conditional deletion of *Atp13a2* in non-human primates, dogs, rats, and mice induces prominent microgliosis and astrogliosis, followed by motor and non-motor behavioural abnormalities, although the extent of dopaminergic neuron loss varies across models^1,2,20–22^. Microglia, the resident innate immune cells of the brain, are highly dependent on lysosomal and mitochondrial fitness, and their dysfunction can promote neuroinflammation, neurotoxicity, and behavioural impairment in PD and related neurodegenerative disorders^23–26^. These observations raise the possibility that microgliosis in *Atp13a2*-deficient models may not be merely a secondary response to neuronal injury, but an early and potentially targetable consequence of disrupted polyamine handling.

Here, we investigated how polyamine dysregulation contributes to gliosis and behavioural dysfunction in *Atp13a2*-deficient mice. We show that brain polyamine deficiency emerges early in disease progression and that further polyamine depletion exacerbates parkinsonian behavioural abnormalities. Conversely, oral Spd supplementation restores brain polyamine availability, attenuates inflammatory microglial activation, and improves motor and non-motor phenotypes. Mechanistically, Spd uptake in *Atp13a2*-deficient microglia counteracts lysosomal dysfunction, mitochondrial oxidative stress, and inflammatory activation, which depends on the the related polyamine transporter Atp13a3. Protective effects of Spd supplementation have also been observed in human iPSC-derived and *Drosophila melanogaster* models of parkinsonism. Together, we identify brain polyamine deficiency as a reversible and therapeutically targetable contributor to neuroinflammation and parkinsonian dysfunction caused by ATP13A2 loss.

## Material and methods

### All protocols and materials are listed in Supplementary Table 1. Postmortem histological examination

The brain underwent neuropathological examination to confirm the diagnosis of ATP13A2-deficiency and exclude other neuropathological alterations^27^. Briefly, formalin-fixed paraffin-embedded tissue (FFPE) samples from neocortical regions, the amygdala, the brainstem, the limbic system and the cerebellum were processed and stained according to current neuropathological neurodegeneration grading schemes^28,29^.

Representative histological images from the frontal cortex (gyrus frontalis superior), the midbrain including the substantia nigra and the striatum are shown. For this, haematoxylin and eosin (HE) was performed using standard laboratory procedures. Immunohistochemical staining was achieved using a Ventana Benchmark XT Autostainer (Ventana, Tucson, AZ, USA), in accordance with the manufacturer’s recommendations, using antibodies against human glial fibrillary acidic protein (GFAP; 1:200, Agilent clone 6F2, M0761, RRID:AB_2109952), Iba-1 (1:200, rabbit polyclonal antibody, synaptic systems, 234 003, RRID:AB_10641962) and CD68 (1:200, Agilent, GA60961-2, RRID:AB_2661840). Slides were electronically scanned at high magnification (×40) as high-resolution images (1900× 1200 pixels) with a NanoZoomer 2.0-HT (Hamamatsu Photonics, Hamamatsu, Japan).

### Mice and genotyping

Mice were generated as previously described ^2^,housed and bred under a 12:12 hour light/dark photocycle with access to water and food *ad libitum.* All protocols and procedures were approved by the Bioethical Committee of the KU Leuven (Project number P069-2021) and conformed to the guidelines of the European Communities Council Directive of November 24, 1986 (86/609/EEC).

*Atp13a2* wildtype (WT) and KO mice were generated by crossing *Atp13a2* heterozygous males and females on a C57/BL6J background. To generate the *Atp13a2* heterozygous mice, *Atp13a2*-Flox male mice with LoxP sites surrounding exon 2 and 3 (RRID: IMSR_JAX:028387) were crossed with female mice expressing Sox2-cre (RRID: IMSR_JAX:008454) to excise the floxed sites of *Atp13a2*. A randomised, equal numbers of male to female WT and KO mice were included across all experiments. The number of mice per experiment was pre-determined based on previous studies assessing the *Atp13a2* KO mouse model ^2^. All experiments were conducted with the researcher blinded to the genotype of the animals.

To validate *Atp13a2* KO and determine genotypes, DNA was extracted from the tail tips of postnatal day (PND)2 mice were collected for polymerase chain reaction (PCR). Two sets of primers were utilised with the following sequences, Floxed/WT/KO (Forward: 5’-CTG CAG CTT CGA GAG GAA AG-3’), WT/Floxed (Reverse: 5’-CAC TCT GTC CTC AGG CCT TC-3’), and KO (Reverse: 5’-TGA GAA GTG GGA ATC GGG-3’). The amplified products produced bands at 426 bp for *Atp13a2* WT and 281 bp for *Atp13a2* KO when visualised using Midori Green on a 1.5% agarose gel.

### Polyamine modulation and supplementation strategies

*Atp13a2* KO and WT mice were treated with negative or positive polyamine modulators depicted in (Fig. 1a) starting at post-natal day (PND)2 to assess the impact of polyamine modulation on mice survival and phenotype. Body weight, food and water intake were monitored over the course of all experiments.

**Figure 1.**
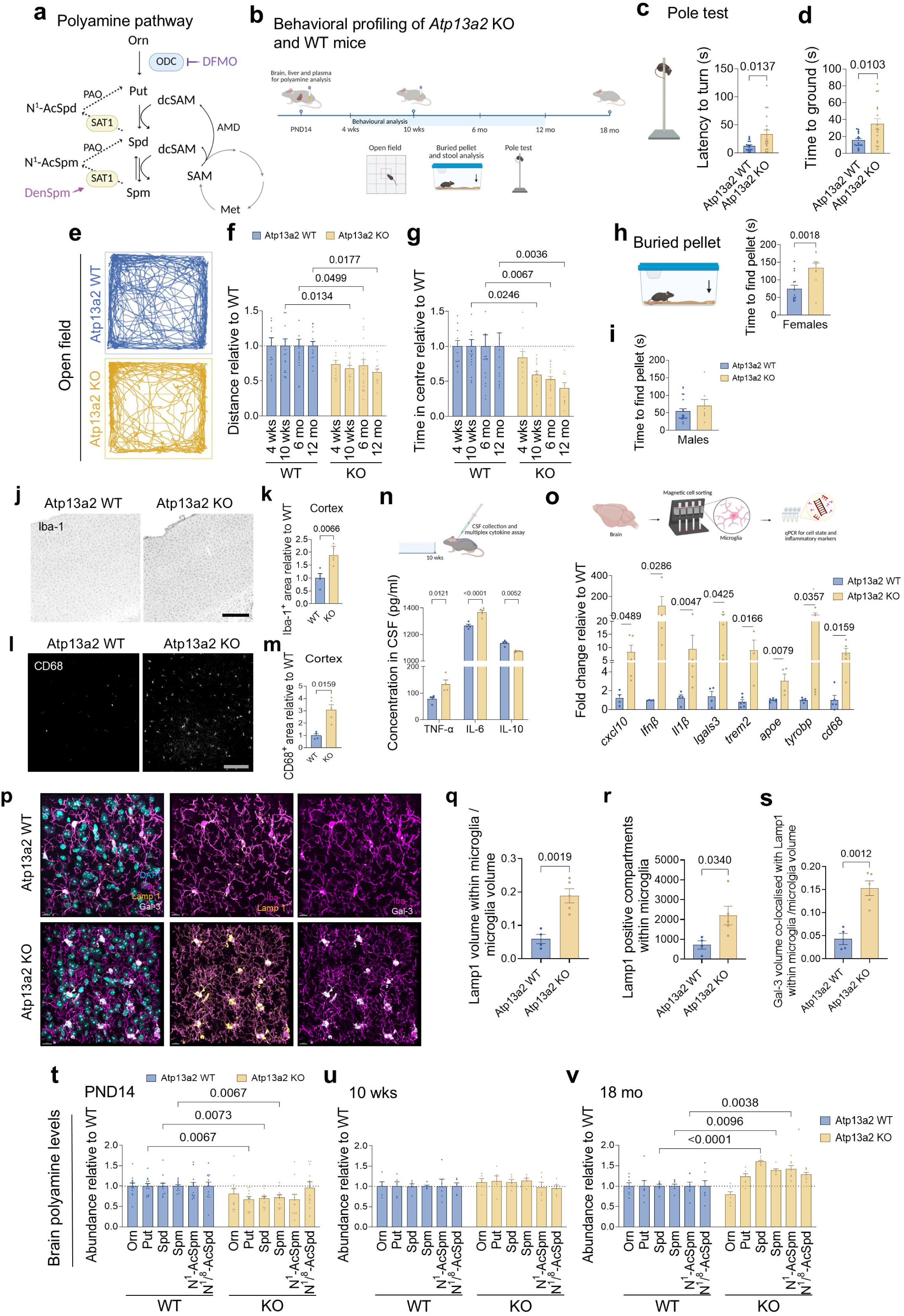
Atp13a2 loss drives early polyamine deficiency preceding the development of neuroinflammation and parkinsonian deficits. (a) Schematic depicting polyamine synthesis and catabolism pathway. (b) Timeline for profiling of behavioural and polyamine signatures in the *Atp13a2* KO and WT mice. (c) Illustration of pole test for mice and latency to turn *Atp13a2* WT and KO mice in the pole test at 10 weeks of age. N = 16 per genotype (d) Time to reach the ground in the pole test at 10 weeks of age. N = 16 per genotype. (e) Representative open field tracks depicting the activity patterns of *Atp13a2* WT and KO mice at 10 weeks of age. (f) Total distance travelled by *Atp13a2* WT and KO mice in the open field at 4 weeks, 10 weeks, 6 and 12 months. 4 weeks, N = 12 WT and 11 KO mice; 10 weeks, N = 19 WT and 17 KO mice; 6 months, n = 20 per genotype. 12 months, N = 13 WT and 11 KO mice. (g) Time spent in the centre of the open field. 4 weeks, N = 12 WT and 11 KO mice; 10 weeks, N = 19 WT and 17 KO mice; 6 months, N = 20 per genotype. 12 months, N = 13 WT and 11 KO mice. (h) Schematic illustrating the buried pellet test and time for female *Atp13a2* WT and KO mice to locate the hidden pellet in the buried pellet test at 10 weeks of age. N = 10 per genotype. (i) Total time in the buried pellet for 10-week-old male mice. N = 11 WT and 7 KO mice. (j) Representative images of cortical microglia (Iba-1, black) of *Atp13a2* WT and KO mice. Scale bar = 200 µm. (k) The area positive for Iba-1 in the cortex. N = 4 WT and 5 KO mice. (l) Representative images of cortical microglia (CD68, white) of *Atp13a2* WT and KO mice. (m) The area positive for CD68 in the cortex. N = 4 WT and 5 KO mice. (n) Cytokine concentrations in the CSF of 10-week-old *Atp13a2* WT and KO mice. N = 4 per genotype. (o) Isolation and assessment of gene expression of a focused subset of pro-inflammatory and disease-associated genes in microglia from 10-weeks-old *Atp13a2* WT and KO mice. N = 4 per genotype. (p) Representative images of Lamp1 (yellow) and Galectin-3 (Gal-3,white) within striatal microglia (Iba-1, magenta) of *Atp13a2* WT and KO mice. Scale bar = 10 µm (q) Lamp1 volume within microglia per microglia volume. N = 4 WT and 5 KO mice. (r) Number of Lamp1 positive compartments within microglia. N = 4 WT and 5 KO mice. (s) Volume of Gal-3 co-localised with Lamp1 per microglia volume. N = 4 WT and 5 KO mice. (t) Abundance of polyamine species within the brains of PND14 *Atp13a2* WT and KO mice. N = 12 WT and 11 KO mice. (u) Abundance of polyamines in the brain at 10 weeks. N = 5 WT and 6 KO mice. (v) Abundance of polyamines in the brain at 18 months of age. N = 7 per genotype.. Data are represented as mean ± SEM. Mann-Whitney test (c-d). Two-way ANOVA with Bonferroni multiple comparisons post-test (f-g). Unpaired Student’s t-test (h). Mann-Whitney test (m). Mixed effects analysis with Bonferroni multiple comparisons post-test (n-o). Unpaired Student’s test (q-s). Mixed effects analysis with Šídák’s multiple comparisons post-test (t-v). Ornithine, Orn; Putrescine, Put; Spermidine, Spd; Spermine, Spm; N^1^-Acetylated Spermine, N^1^-AcSpm; N^1^/N^8^-Acetylated Spermidine, N^1^/N^8^-AcSpd. Illustrations were made on Biorender.com.

α-difluoromethylornithine (DFMO; a kind gift from Dr. Patrick Woster’s lab at the University of South Carolina, and CoreRX, Clearwater, FL) was administered intraperitoneal (*i.p.*) at a dose of 0.35 mg/g/day on PND2-5. The dose was then adjusted according to body weight to 0.4 mg/g/day on PND6-14, and to 0.5 mg/g/day on PND15-21. After weaning, mice received DFMO via drinking water (20 mg/ml, average daily intake of 1.6 g/kg/day) until 8 weeks of age. N^1^, N^11^-Diethylnorspermine tetrahydrochloride (DenSpm; Torcis, 0468) was administered *i.p.* at a dose of 0.06 mg/g/day from PND2 until 4 weeks of age. Mice also received *i.p.* injections of Spd (0.1 mg/g/day; Acros organics, 215100250) or Spm (0.05 mg/g/day; Fisher Scientific, J63060) from PND2 to PND14.

For polyamine supplementation, a cohort of *Atp13a2* KO and WT mice were supplemented with Spd or Spm from PND2 to 10 weeks of age, whereas a second cohort received Spd supplementation from 7 to 12 months of age. During early stages of development, mice received polyamine supplementation through the lactating mothers that received drinking water supplemented with 3 mM of Spm, Spd or control water. After weaning and until 10 weeks of age, mice received Spd (3 mM, 0.764 mg/ml, average 75 mg/kg/day) or Spm (3 mM, average 117 mg/kg/day) through drinking water as previously described ^30–32^. Similarly, mice aged 7 months received Spd (3 mM, 0.764 mg/ml) through drinking water until 12 months of age. Oral Spd and Spm effects were all cross compared to one control cohort assessed in the same experiment.

### Plasma and tissue collection

Plasma of PND14 and 10-week-old *Atp13a2* KO and WT mice was analysed by targeted metabolomics to assess polyamine levels. To collect plasma, blood was drawn from the facial veins of PND14 and 10-week-old *Atp13a2* KO and WT mice, and collected in EDTA coated tubes (Sarstedt, 20.1341). To separate the plasma, blood samples were centrifuged at 2,000 x g for 15 minutes (min) at 4°C. Plasma was then stored at −80°C.

The livers (PND14) and brains (PND14, 10 weeks and 18 months old) of *Atp13a2* KO and WT mice were collected for metabolomics analysis. The tissue was dissected out following cervical dislocation, snap-frozen in liquid nitrogen and stored at −80°C until used.

The brains of 10-week-old *Atp13a2* KO and WT were collected following transcardial perfusion and later processed for immunohistochemistry. Briefly, mice were given a lethal dose of sodium pentobarbital (Dolithol *i.p*. at 100 mg/kg) and transcardially perfused with 0.9% saline followed by 4% paraformaldehyde (PFA; Sigma-Aldrich, 441244). Brains were dissected and postfixed overnight in 4% PFA. Sagittal brain sections were then cut on a vibrating microtome at a thickness of 40 µm and processed for immunohistochemistry. A subset of mice was transcardially perfused with 0.9% saline, and the brains were processed for magnetic cell sorting for isolating microglia (see below).

### Cerebrospinal fluid collection

CSF was collected through a cisterna magna puncture to assess inflammatory markers in the 10-week-old *Atp13a2* KO and WT mice as previously described ^33^. Mice were first anaesthetised and secured in a stereotaxic frame. To expose the cisterna magna, the dorsal neck area was shaved, sterilised and the overlying muscles retracted. A pulled glass capillary mounted on a micromanipulator was inserted at a 45° angle into the cisterna magna, avoiding blood vessels. CSF was collected by capillary action, centrifuged at 10,000 x g for 10 min at 4°C, and stored at −80°C for later assessment of cytokine levels.

### Motor and non-motor behavioural tests

*Atp13a2* KO and WT mice were tested for motor and non-motor functions at 4 weeks, 10 weeks, 6 months and 12 months of age (n = 11-20 per genotype for each age group). All mice were tested during the light phase and were habituated for 30 min in the testing room prior to testing in the following paradigms.

#### Open field

*Atp13a2* KO and WT mice were evaluated for locomotion and exploratory behaviour using the open field test as described previously ^34^. *Atp13a2* KO and WT mice activity was recorded for 30 min using an overhead camera and analysed using ANYmaze software (RRID:SCR_014289). Parameters measured included distance, immobility, freezing and time spent in the periphery and the centre.

#### Pole test

The balance and coordination of *Atp13a2* KO and WT mice was evaluated with the pole test ^35^. Briefly, *Atp13a2* KO and WT mice were positioned at the top of a pole (50 cm in height and 0.8 cm in diameter) placed in an empty cage with the head orientated upwards. Time taken for mice to reorientate downwards (time to turn) and the time taken to reach the cage (total time) were recorded.

#### Limb clasping

Motor-deficits of *Atp13a2* KO and WT mice were also evaluated by the limb clasping test ^36^. Over two trials, *Atp13a2* KO and WT mice were suspended by the tail for 10 seconds (s) with a 5-10 s interval between trials. A score from 0 to 4 was given based on the severity of clasping behaviour. Animals received a score of 0 when the hind limbs were consistently splayed outwards, away from the abdomen. A score of 1 was given when one of the hind limbs was retracted towards the abdomen for more than 50 % of the time suspended. A score of 2 was given when the two hind limbs were partially retracted toward the abdomen for more than 50% of the time suspended. A score of 3 was given when the hind limbs were entirely retracted and touching the abdomen for more than 50% of the time suspended. A score of 4 was selected when the fore and hind limbs were retracted for more than 50% of the time suspended.

#### Buried pellet

With the buried pellet test we assessed olfactory dysfunction in *Atp13a2* KO and WT mice as previously described ^37^. *Atp13a2* KO and WT mice were food deprived for 20 hours (h) prior to testing. The time required for mice to locate a pellet buried under a 3 cm layer of odour-neutral bedding was recorded (cutoff at 300 s). Mice were offered two trials to locate the buried pellet over two non-consecutive days.

#### Stool count and moisture content

Stool produced by *Atp13a2* KO and WT mice was collected by placing each mouse individually in an empty cage for 60 min. Stools were counted and collected to calculate the moisture content ^38^. Moisture content was determined as the difference in weight of stools prior to and following drying in an oven at 65°C for 18 h.

### Metabolomics

Plasma (PND14 and 10 weeks), brain (PND14, 10 weeks and 18 months) and liver (PND14) samples of *Atp13a2* KO and WT mice were analysed to detect relative values of different polyamine species (Ornithine, Orn; Putrescine, Put; Spermidine, Spd; Spermine, Spm; N^1^-Acetylated Spermine, N^1^-AcSpm; N^1^/N^8^-Acetylated Spermidine, N^1^/N^8^-AcSpd).

Metabolomics was performed by the VIB metabolomics core (Dr. B. Ghesquire, KU Leuven). Briefly, plasma samples or brain tissue diluted in 80 % methanol were processed through a liquid chromatography-mass spectrometry (LC-MS) using a Dionex UltiMate 3000 LC System (Thermo Fisher Scientific) coupled to a Q Exactive Orbitrap mass spectrometer (Thermo Fisher Scientific) operated in positive mode. Chromatographic separation was performed using Poroshell 120 HILIC-Z PEEK column (PEEK, 2.1 × 100 mm, 2.7 µm; Agilent InfinityLab). During LC, a linear gradient was carried out with acetonitrile (Solvent A) and ammonium formate (10 mM, pH 3.8, solvent B) under a constant temperature of 25°C. MS was operated at full scan (Range 70-150) with a spray voltage of 3.2 kV, a temperature of 320°C, sheath gas at 40.00 and auxiliary gas at 10.0. The AGC target was set at 3 x 10^6^ using a 70,000 resolution. Data was collected with Xcalibur software (Thermo Fisher Scientific) and analysed by the integration of peak areas.

### Immunohistochemistry

#### Chromagen labelling

Sagittal brain sections from *Atp13a2* KO and WT mice were first washed in phosphate buffered saline (PBS), then incubated in an antigen retrieval buffer (1 M citrate buffer, pH 6) for 30 min at 80°C and an additional 20 min on ice. Tissue sections were then washed 3 times in PBS and incubated in a blocking solution (3% H2O2 and 10% methanol in PBS) for 10 min at room temperature. After washing in PBS, sections were incubated overnight at room temperature with goat anti-Iba-1 (1:500; Abcam, ab5076, RRID: AB_2224402) or rabbit anti-GFAP (1:2000; Agilent, z0334, RRID: AB_10013382). Tissue sections were subsequently incubated with a biotinylated rabbit anti-goat (1:600; Agilent, E0466, RRID:AB_3676677) or a biotinylated goat anti-rabbit (1:600; Abcam, ab6720, RRID: AB_954902) for 30 min at room temperature. Sections were then incubated for 30 min at room temperature with HRP-conjugated Streptavidin (1:1000; Agilent, P0397). Immunolabelling was revealed with 3,3’-Diaminobenzidine tetrahydrochloride solution (DAB, 40% in PBS with 0.028% H_2_O_2,_ Sigma, D5905). Sections were afterwards mounted on pre-coated slides and dehydrated in ascending concentrations of ethanol. Tissue sections were next cover-slipped using DPX (Sigma-Aldrich, 06522) as mounting media.

#### Immunofluorescent labelling

Brain tissue was washed in PBS then incubated for 40 min in a blocking solution containing 10 % donkey serum (Jackson Immuno Research, 017-000-121). Sections were next incubated overnight at room temperature with the primary antibody, rabbit anti-CD68 (1:500; Abcam, ab125212, RRID: AB_10975465), mouse anti-Galectin-3 (Gal-3; 1:500; Abcam, ab2785, RRID:AB_303298), rat anti-Lamp1 (1:500; Santa Cruz Biotechnology, sc-19992, RRID:AB_2134495), mouse anti-RELA/NF-κB p65 (1:500; Santa Cruz Biotechnology, Sc-8008, RRID: AB_628017) and goat anti-Iba-1 (1:500) in blocking solution. Tissue sections were washed in PBS and incubated afterwards with donkey anti-rabbit Alexa 488 (1:500; Abcam, ab150073, RRID:AB_2636877), goat anti mouse Alexa 546 (1:500; Thermo Fisher Scientific, A-11003, RRID:AB_2534071 Goat anti Rabbit Alexa 488 (1:500; Thermo Fisher Scientific, A-11008, RRID:AB_143165), Goat anti rat Alexa 647 (1:500; Thermo Fisher Scientific, A-21247, RRID:AB_141778) for 2 h at room temperature. Sections were mounted on slides and counterstained with DAPI (1:500; MedChemExpress, HY-D0814).

#### Image acquisition and analysis

Brain sections immunolabelled for Iba-1 and GFAP (chromogen) were imaged using the Leica aperio CS2 slide scanner at 20x magnification. Sections immunolabelled for CD68 were imaged using the Leica DM6 B LED fluorescent microscope at 40x magnification. Using ImageJ (RRID:SCR_003070; NIH, Bethesda, MD, USA), the cortex, striatum and cerebellum were assessed for the percentage area positive for Iba-1, GFAP or CD68.

Brain sections immunolabelled for Gal-3, RELA/NF-κB p65, Lamp1, Iba-1 were imaged using LSM 880 Airyscan confocal microscope at 40x magnification (1.3 Oil) with z-stack images were captured at a focal density of 0.48 µm with a pinhole of 1 airy unit (AU), and averaging of 2. Images were analysed using Imaris software (RRID:SCR_007370).

Surface creation was used to 3D model microglia using Iba-1 signal, then Lamp1 signal was masked onto the Iba-1 positive surface and a surface for Lamp1 was created from the masked channel. Gal-3 signal was then masked onto the Lamp1 and a surface was thereafter created for the Gal-3 mask. Volumes of all surfaces were measured and Lamp-1 and Gal-3 surface volumes normalised to Iba-1 surface volume. Intensity thresholds and parameters used to create the surfaces were based on negative control lacking primary antibodies and were fixed across all images.

### Isolation of microglia via magnetic cell sorting

Microglia were isolated from the cortices of *Atp13a2* KO and WT mice with and without Spd oral supplementation as previously described ^39^. Tissue was enzymatically dissociated in HBSS with collagenase A and DNase I at 37°C for 15 min, then mechanically dissociated with a flame-polished glass pipette. The resulting single-cell suspension was filtered through a 70 µm filter then centrifuged for 10 min at 300 x g at 4°C. Cellular debris was removed through a gradient with a debris removal solution, cells were then washed in PBS and centrifuged for 10 min at 1,000 x g at 4°C. Cells were then resuspended in MACS buffer (PBS with 1 mM EDTA, Thermo Fisher Scientific, AM9261; 1% bovine serum albumin, BSA, Carl Roth, 8076.4) and incubated with anti-CD11b magnetic microbeads for 15 min on ice. After washing the cells in MACS buffer, CD11b-positive cells were enriched through magnetic columns according to manufacturer instructions. Isolated microglia were collected in a guanidine-thiocyanate containing lysis buffer (RLT, Qiagen, 74004) and stored at −80°C until processing for qPCR.

### Primary microglia and neuronal cultures

Microglia and midbrain neurons were isolated from PND0-2 *Atp13a2* KO and WT as previously described ^40,41^. Briefly, *Atp13a2* KO and WT pups were decapitated and brains dissected out in HBSS (Life technologies, 14175079; Supplemented with 10 mM HEPES, Gibco, 15630080). The meninges were carefully removed and brain tissue collected in DMEM 10:10:1 (Gibco, 11965092; supplemented with 10% horse serum, Gibco, 2605-088; 10% foetal bovine serum, FBS, Gibco, A5209501; 1% Penicillin/streptomycin, Sigma, P4458) for microglia cultures, and HBSS supplemented with 10 mM HEPES for neuronal cultures.

Brain tissue was dissociated to a single cell suspension and plated in poly-D-lysine coated (PDL; Life technologies, A3890401) Ti75 flasks to generate mixed glia cultures. Cultures were gently shaken at 0.42 x g for 2 h to isolate microglia from the underlying glia cells layer on day *in vitro* (DIV)10. Microglia were then seeded on 5 µg/ml PDL coated coverslips in 12-well plates at a density of 2 x 10^5^ cells per well. Neurons were seeded at a density of 2 x 10^4^ cells per well in 24-well plates coated with PDL and laminin (1 µg/ml). Microglia were maintained in DMEM 10:10:1, whereas neurons were maintained in neurobasal medium (Supplemented with 2% B-27, Life technologies, 17504044; 2% glutamax, Invitrogen, 35050061; 1% Penicillin/streptomycin) at 5% CO_2_ and 37°C.

#### Pharmacological modulation and Atp13a3 knockdown

To model inflammation in microglia, microglia were treated with 100 ng/ml lipopolysaccharide (LPS; Sigma-Aldrich, serotype 0111:B4) in DMEM 10:10:1 for 24 h. The anti-inflammatory effects of Spd (0.01 and 0.3 µM) were assessed in combination with pharmacological modulators, such as the polyamine transport inhibitor AMXT 1501 (1 µM pretreatment for 1 h prior to LPS), and the mitochondrial antioxidant MitoTEMPO (1 µM; Sigma, SML0737). Spd and AMXT 1501 treatment was performed in combination with 1 mM of aminoguanidine to inhibit polyamine oxidases in the sera ^42^. Following incubation times, microglia were fixed in 4% PFA) for 20 min then processed for immunocytochemistry or collected in a guanidine-thiocyanate containing lysing buffer, snap-frozen in liquid nitrogen and stored at −80°C for qPCR. Media supernatant was spun at 300 x g for 10 min at 4°C then stored at −80°C for cytokine analysis.

For establishing the effects of Spd treatment on neuronal cultures, microglia were treated with 0.3 µM Spd for 2 h, shaken at 0.42 x g for 2 h and resuspended in 50:50 ratio of neurobasal medium and DMEM 10:10:1. Microglia were then added in 1:4 ratio to neuronal cultures on DIV11. Co-cultures were maintained for 72 h then fixed with 4% PFA for 10 min on ice.

*Atp13a3* knockdown was performed using ON-TARGETplus SMARTpool siRNA (Horizon, L-051693-01-0010) delivered in DharmaFECT transfection reagent (Horizon, T-2005-01) following the instructions of the manufacturer. The *Atp13a3* siRNA target sequences are 5’-GAAGAAAAUAGAUGCGAGA-3’, 5’-CUGCCUGAGUGGCGAGUGA-3’, 5’-GAUACAUGGCUAUAACUUA-3’, and 5’-UAUCAACAAGGGUCAAGAA-3’, and non-targeting siRNA, 5’-UGGUUUACAUGUCGACUAA-3’, as control. OptiMEM (64 µl; Gibco, 31985062) and siRNA (5, 10 or 100 nM) were mixed with 2 µl of DharmaFECT transfection reagent in delivery media in a total volume of 250 µl, incubated for 30 min at room temperature and then added to each well at 5% CO_2_ and 37°C. After 24 h, microglia were treated with Spd and LPS, and fixed in 4% PFA for immunocytochemistry. For qPCR, microglia were resuspended in a guanidine-thiocyanate containing lysis buffer then snap-frozen in liquid nitrogen and stored at −80°C.

#### Immunocytochemistry and image analysis

Cover slips were washed 3 times in PBS and incubated in a blocking solution (1% FBS, and 0.1% BSA) for 1 h at room temperature. Cells were then incubated with rabbit anti-Iba-1 (1:1000; WAKO, 019-19741, RRID:AB_839504) or chicken anti-MAP2 (1:500; Abcam, ab5392, RRID:AB_2138153) in blocking solution overnight at 4°C. Cells were washed in PBS then incubated with goat anti-rabbit 488 (1:500; Thermo Fisher Scientific, R37116, RRID:AB_2556544) or goat anti-chicken 488 (1:500; Invitrogen, A32931TR, RRID:AB_2866499) for 2 h at room temperature. DAPI was applied to visualise nuclei. Mitochondrial reactive oxygen species (MitoROS) were detected by MitoSox (1 µM; Thermo Fisher Scientific, M36008) in live cells.

Fluorescent images were captured with the LSM880 Airyscan confocal at 40x magnification, z-stack images were captured at a focal density of 0.43 µm with a pinhole of 1 airy unit (AU), and averaging of 2. Images were analysed using Image J software (RRID:SCR_003070; NIH, Bethesda, MD, USA). Microglia aspect ratio was assessed to evaluate cell shape and morphology (aspect ratio values of 1 reflect circular cells, while higher values indicating enhanced ramification). Intensity of staining was also analysed as the mean intensity or integrated density (average grey values and area product). Neurite length was measured using the simple neurite plugin in Image J ^43^.

Independent biological replicates were generated from a minimum of three independent litters per genotype (5-8 pups per litter). The averages of ≥ 20 cells are reported for two or more independent biological replicates. Biological replicates are represented as individual data points.

### Cytokine analysis

Cytokine levels were determined in the CSF and media supernatant using a V-PLEX Proinflammatory Panel 1 Mouse Kit (K15048D, Meso Scale Discovery) following the manufacturer instructions. Samples and standards were incubated in wells pre-coated with analyte-specific capture antibodies (including TNF-α, IL-6 and IL-10) then detected with SULFO-TAG-conjugated secondary antibodies. Electrochemiluminescent signal was detected with MESO QuickPlex SQ 120MM (Meso Scale Discovery).

### RNA extraction and qPCR

Total RNA was extracted using the RNeasy micro kit (Qiagen, 74004) following the manufacturer’s protocol. The RNA concentration and purity were assessed using bioanalyzer. For cDNA synthesis, 0.5 µg of total RNA was reverse-transcribed using the iScript cDNA Synthesis kit (Biorad, 1708891) according to the manufacturer’s instructions. The resulting cDNA was diluted 1:10. qPCR was performed with the powerup SYBR master mix (Thermo Fisher Scientific, A25742) on Applied Biosystems QuantStudio 6 Pro Real Time PCR system (Thermo Fisher Scientific, RRID:SCR_028138). The following primers were utilised: *Atp13a3* (Forward: 5’-ATGGCACCGGATCAGAAGAC-3’, Reverse: 5’-ATCACCGCACATCCCAACAA-3’), and *gapdh* (Forward: 5’-CCTGCTTCACCACCTTCTTGA-3’, Reverse: 5’-TGTGTCCGTCGTGGATCTGA-3’). Relative gene expression was analysed using the ΔΔCt method, normalising Atp13a3 expression to Gapdh.

Primers used include: *trem2* (Forward: 5’-AACTTCAGATCCTCACTGGACC-3’, Reverse: 5’-TCCTGCTCCCAGGATAGGTG-3’). *cd68* (Forward: 5’-GGGGCTCTTGGGAACTACAC-3’, Reverse: 5’-GTACCGTCACAACCTCCCTG-3’). *Il-1b* (Forward: 5’-CTGCAGCTGGAGAGTGTGG-3’, Reverse: 5’-GGGGAACTCTGCAGACTCAA-3’). *Ifnb* (Forward: 5’-CTCCAGCTCCAAGAAAGGAC-3’), Reverse: 5’-TGGCAAAGGCAGTGTAACTC-3’). *Cxcl10* (Forward: 5’-AAGTGCTGCCGTCATTTTCT-3’, Reverse: 5’-GTGGCAATGATCTCAACACG-3’). *Lgals3* (Forward: 5’-CCCAACGCAAACAGGATTGT-3’, Reverse: 5’-GAAGCGGGGGTTAAAGTGGA-3’). *Tyrobp* (Forward: 5’-*CTCCTGACTGTGGGAGGATTAAG*-3’, Reverse: 5’-*ACAATCCCAGCCAGTACACC*-3’). *Apoe* (Forward: 5’-CACAAGAACTGACGGCACTG-3’, Reverse: 5’-TGCGTAGATCCTCCATGTCG-3’).

### iPSC Culture

Human induced pluripotent stem cell (iPSC) lines derived from a male donor (KOLF2.1J background), including the ATP13A2^c.1306^ mutant line and its isogenic parental control (ATP13A2^WT^), were obtained from the iPSC Neurodegenerative Disease Initiative (iNDI; generously provided by Bill Skarnes).

Cells were maintained at 37°C in a humidified atmosphere containing 5% CO₂. Cultures were grown on Geltrex-coated plates (Thermo Fisher Scientific, A1413201) in StemFlex medium (Gibco, A3349401). Upon reaching approximately 70-80% confluence, cells were passaged using Accutase (STEMCELL Technologies, 07920) for enzymatic dissociation and replated in fresh StemFlex medium. To enhance survival following single-cell dissociation, 10 µM Y-27632 ROCK inhibitor (Tocris, 1254) was added to the culture medium for the first 24 hours after replating.

### Differentiation of iPSCs into Microglia

To generate iPSC-derived microglia (iMG), 2.5 × 10⁶ human iPSCs were seeded into AggreWell™ 24-well plates (STEMCELL Technologies, 34811) to form embryoid bodies (EBs). Prior to cell seeding, wells were pretreated with 500 µL Anti-Adherence Rinsing Solution (STEMCELL Technologies, 07010) and centrifuged at 1300 × g for 5 min to facilitate efficient EB recovery the following day. Cells were plated in 2 mL StemFlex™ medium supplemented with 10 µM Y-27632 (day 0) and centrifuged at 100 × g for 3 min to promote uniform aggregation.

On day 1, approximately 20 EBs per well were transferred to Matrigel-coated 6-well plates and maintained in StemFlex™ medium containing 1% penicillin–streptomycin. Hematopoietic differentiation was initiated on day 2 by replacing the medium with STEMdiff™ Hematopoietic Kit Medium A (2 mL/well). Medium A was replenished on day 4, followed by a switch to Medium B on day 5 to support hematopoietic progenitor cell (HPC) maturation.

Floating HPCs were harvested on days 11 and 13 and replated onto Matrigel-coated 6-well plates. Cells were then differentiated toward a microglial lineage in microglia differentiation medium consisting of DMEM/F12 (Gibco, 11320-033) supplemented with 2× B27 (Thermo Fisher Scientific, 17504044), 0.5× N2 (Thermo Fisher Scientific, 17502048), 1× GlutaMAX (Gibco, 35050061), 1× non-essential amino acids (Gibco, 11140-050), 400 µM monothioglycerol (MilliporeSigma, M6145), and 5 µg/mL human insulin (MilliporeSigma, I9278). Media were freshly supplemented with 100 ng/mL IL-34 (PeproTech, 200-34), 50 ng/mL TGF-β1 (PeproTech, 100-21), 25 ng/mL M-CSF (PeproTech, 300-25), and GM-CSF (PeproTech, 300-03; concentration as indicated). Cells were maintained under these conditions for 25-28 days to allow microglia maturation.

Beginning on day 20 and continuing through full iMG maturation, cultures were additionally supplemented with 100 ng/mL CD200 (Thermo Fisher Scientific, 310-46) and 100 ng/mL CX3CL1 (PeproTech, 300-3) to promote microglial homeostatic phenotypes.

For Spd supplementation experiments, Spd (Millipore Sigma, S2626) was diluted into media at 0.3 µM and incubated for 2 h before fixing.

### Immunofluorescence and Confocal Microscopy

Microglia cells were seeded in 96-well black-walled plates (Greiner, 655090) at 30,000 cells per well on day 22-25 and were processed for staining fixed with 4% PFA for 15 min at room temperature. Permeabilisation and blocking were performed for 1 h at room temperature in a buffer consisting of 0.3% Triton X-100 and 5% donkey serum in PBS. Primary antibodies (Iba-1, WAKO, 019-19741, RRID:AB_839504; CD68, Abcam, AB955, RRID:AB_2138153) was diluted 1:300 in blocking buffer and incubated overnight at 4°C. Following three PBS washes, wells were incubated with species-specific Alexa Fluor-conjugated secondary antibodies at 1:500 for 1 h at room temperature. Nuclei were counterstained with Hoechst 33342 (Cayman Chemical, 13197) for 5 min.

### CellROX Green oxidative stress imaging (live staining with post-fixation imaging)

Cells were incubated with CellROX Green Reagent (Thermo Fisher Scientific, C10444) dilute in complete culture medium at 1µM final concentration for 30 min at 37°C protected from light. After staining, the dye-containing medium was removed and replaced with fresh medium. Cells were then immediately imaged using GFP/FITC channel (excitation/emission ∼485/520 nm).

### Microscopy imaging and analysis

Confocal immunofluorescence images were obtained using either a Nikon Ti2-E AX confocal microscope or a High Content Imager CX7 (Thermo Fisher) operated with HCS Studio 4.0. For histological evaluation, immunohistochemistry (IHC) slides were imaged at 40× magnification using an Aperio VERSA 8 slide scanner (Leica Biosystems, Wetzlar, Germany). Representative regions of interest were selected and imaged with Aperio ImageScope software (Leica Biosystems). To maintain consistency across experiments, all images were acquired using identical laser power, gain, and exposure parameters.

CellProfiler Image Analysis Software (RRID:SCR_007358) was employed to automate image analysis using pipelines for cell counting, area-based thresholding, and quantification of signal intensity^44^. The software was specifically used to segment nuclei and define cellular boundaries, enabling determination of the number of cells positive for specific markers as well as the total stained area.

## Drosophila experiments

### Fly stocks and rearing conditions

*Drosophila melanogaster* fly lines were generated as described previously using CRISPR/Cas9-mediated gene editing ^45^. The following fly genotypes were examined, along with their stock and abbreviation. Genotype: w[1118] M ^40^; stock: Dmel\Canton-S-iso-M{w+} w1118 (abbreviated: CSw1118w^+^). Genotype: w[1118]; TI{w[+]=white-STAR}anne[KO-WS]/ci[D]; stock: Dmel\Canton-S-iso-TI{white-STAR}anne^KO-WS^ (abbreviated: anne*^KO-WS/+^*). Genotype: w[1118] TI{w[+]=white-STAR}Pink1[KO-WS]/FM7a; stock: Dmel\Canton-S-iso-TI{white-STAR}Pink1^KO-WS^ (abbreviated: Pink1^KO-WS/y^). Genotype: w[1118]; TI{w[+]=white-STAR2}iPLA2-VIA[KO-WS]/TM6C Tb[1] Sb[1]; stock: Dmel\Canton-S-iso-TI{white-STAR2}^27^iPLA2-VIA^KO-WS^, (abbreviated: iPLA2-VIA^KO-WS^). Genotype: w[1118]; TI{w[+]=white-STAR2}nutcracker[KO-WS]/TM6C Tb[1] Sb[1]; stock: Dmel\Canton-S-iso-TI{white-STAR2}nutcracker^KO-WS^ (abbreviated: nutcracker ^KO-WS^).

Drosophila flies were maintained on a standard cornmeal and molasses diet. For experiments ^45^, flies were raised in parallel on the same batch of food in a temperature- and light-controlled incubator at 25°C and 12-h light-dark cycle, and kept in mixed populations of similar density. Control (CS^w1118^w+) and *anne ^KO-WS/+^* were aged to 42 ± 2 days while *Pink1^KO-WS/y^, nutcracker^KO-WS^* and *iPLA2-VIA^KO-WS^* with their corresponding control were aged to 22 ± 2 days due to their shorter survival. The “/+” refers to heterozygosity and “/y” to hemizygosity.

For Spd treatment, male flies of control and experimental genotypes were collected at 1-3 days post eclosion and aged on treatment food, which was refreshed every two to three days. On the day of the experiment, flies were transferred to fresh treatment food between Zeitgeber (ZT)0 and 1. Treatment food was prepared every one to two weeks and stored at 4°C in darkness. Spd was dissolved in sterile distilled water to make a 500 mM stock solution, aliquoted in single-use portions and stored at −20°C. Spd or sterile distilled water as a vehicle control was added to freshly prepared fly food, heated up to approximately 40°C, to a final concentration of 5 mM. Biological replicates and number of times the experiment was performed is indicated as ‘N’ in the figure legends.

### Startle induced negative geotaxis

Startle induced negative geotaxis (SING) was assessed between ZT 3 and 5 in groups of male flies treated with Spd or solvent control as described previously ^45,46^. Flies aged 22 ± 2 or 42 ± 2 days were transferred without prior anaesthesia into the first tube of the climbing apparatus and allowed to acclimate to the experimental environment (24 ± 1°C, 40-60% humidity) for several minutes. Flies were then tapped down five times and given 30 s to climb. Flies that reached the upper tube were moved to the next tube; the procedure was repeated four times. The number (N) of male flies of each genotype reaching each of the tubes were counted to calculate the SING score. Scores were normalised to the mean SING score of the control.

*SING score = ((*N*_1_ ∗ 0) + (*N*_2_ ∗ 1) + (*N*_3_ ∗ 2) + (*N*_4_ ∗ 3) + (*N*_5_ ∗ 4))/4(*N*_1_ + *N*_2_ + *N*_3_ + *N*_4_ + *N*_5_). *N*_k_* = number of flies in the *k*th tube. Individual data represent groups of animals and N ≥ 9.

### Immunohistochemistry

For immunohistochemistry, 22 ± 2 or 42 ± 2 day old adult brains in at least 2 independent experiments were dissected in ice-cold PBS and fixed for 30 min in freshly prepared 3.7% PFA (in PBX, 1x PBS and 0.2% Triton X-100) at room temperature. After 3 washes in PBX, the brains were incubated in a blocking solution (PBX, 10 % normal goat serum) for 1 h at room temperature. Subsequently, the brains were incubated in primary antibodies (1:200, rabbit anti-TH, Sigma; 1:100, mouse anti-DLG, DSHB) in blocking solution at 4°C for 1.5-2 days. The brains were then washed 3 times in PBX at room temperature. Brains were then incubated with a cocktail of secondary antibodies (1:500, goat anti-rabbit Alexa488 and goat anti-mouse Alexa555) in PBX with 10 % normal goat serum overnight at 4°C. The brains were finally mounted anterior facing up in RapiClear 1.47 (SUNJin Lab) following washes in PBS. Individual data points represent individual animals, N ≥ 18 per genotype.

### Image acquisition and analysis

Focal Z-stacks of whole fly brains were acquired using a Nikon A1R confocal microscope equipped with a 20x water immersion lens (NA0.95). Imaging was performed using a galvano scanner at a zoom factor of 1, scan speed 0.5, and line averaging of 2. All images were acquired with a pinhole of 2.3 AU pinhole and a resolution of 1024 × 1024 pixels. Z-stacks were acquired with 3 µm step intervals, identical image settings were applied across all genotypes and sessions to ensure consistency. Image analysis was performed with ImageJ.

To quantify dopaminergic neuron innervation of the mushroom body (MB), anti-DLG we performed immunostaining to determine the five z-stacks encompassing the synaptic region of the MB lobes. A region of interest (ROI) was defined around the MB in the sum projection of the five z-planes. Within the selected z-stacks, an identical threshold for the anti-TH fluorescence signal was applied for all 5 z-planes to exclude background signals comparable to the control. The area of the threshold within the ROI was quantified in each z-plane, summed, and then normalised to the total MB ROI area for each brain individually (TH+ area/ MB area). For each experiment, the individual TH+ area/MB area values were further normalised to the mean of the control. The maximum age analysed of each genotype is reported as “aged flies”. For representative images the maximum projection of five z-planes as well as the area of the threshold of the middle z-plane is shown.

### ATP13A2 expression analysis in human and mouse

Human RNASeq tissue expression data were downloaded from the GTEX Portal (GTEX V11 Transcripts per Million (TPM) dataset; https://www.gtexportal.org/home/). Mouse RNASeq tissue expression data were downloaded from the MGI Portal (GXD dataset; https://www.informatics.jax.org/). Expression values were converted to log_2_(TPM+1) values and annotated for tissue type based on GTEX or GXD metadata and for system based on general anatomophysiological function.

### Statistical analysis

Statistical analysis was performed using PRISM software version 10.4.1 (GraphPad Prism, RRID: SCR_002798). Age matched *Atp13a2* WT and KO groups were compared using unpaired Students t-test or a Mann-Whitney U-test when comparing one variable. One-way and Two-way ANOVA with multiple comparisons correction were performed to compare age matched *Atp13a2* WT and KO where multiple variables were assessed. Parametric statistical tests were performed when data met statistical assumptions (Using Shapiro-Wilk and D’Agostino-Pearson omnibus for testing normality) otherwise we turned to non-parametric tests. Correction for multiplicity was performed with the false detection rate method.

All values are represented as the mean ± SEM. Specifics on the statistical test are indicated in the figure legends. A *P* value of < 0.05 was considered statistically significant unless corrections for multiplicity using the false detection rate method suggested otherwise. Details of tests used for each graph are reported in the figure legends.

## Results

### Reduced brain polyamine levels precede parkinsonian symptoms in *Atp13a2* KO mice

Loss-of-function mutations in ATP13A2 cause early- or juvenile-onset parkinsonism in humans. As motor deficits in *Atp13a2* KO mice were previously reported to only emerge at ∼9 months of age ^2^, we assessed whether *Atp13a2* deficiency also induces earlier behavioural abnormalities in mice.

Motor and non-motor functions were systematically evaluated in *Atp13a2* KO and WT mice at 4 weeks, 10 weeks, 6 months, and 12 months of age using the open-field assay, pole climbing test, buried food pellet test (Fig. 1b-i, Supplementary Fig. 1a-b). Gastrointestinal function was additionally assessed by measuring faecal pellet output and water content (Supplementary Fig. 1c,d). *Atp13a2* KO mice showed increased latency to turn and descend in the pole test at 10 weeks of age (Fig. 1c,d). Consistent with impaired motor coordination, *Atp13a2* KO mice exhibited a significant reduction in total distance travelled in the open field beginning at 10 weeks of age, which persisted through 6 and 12 months (Fig. 1e,f). This hypo-locomotion was accompanied by increased immobility and freezing behaviour from 10 weeks onwards (Supplementary Fig. 1a,b). Next, we assessed non-motor features associated with parkinsonism ^47^. At 10 weeks of age, KO mice spent less time in the centre of the open field, consistent with increased anxiety-like behaviour (Fig. 1g). At 6 months, female KO mice displayed delayed pellet retrieval in the buried pellet test, indicative of olfactory dysfunction (Fig. 1h,i). In addition, 10-week-old KO mice produced fewer faecal pellets during testing without changes in faecal moisture, suggesting early gastrointestinal abnormalities (Supplementary Fig. 1c,d). Together, these data demonstrate that both motor and non-motor phenotypes are detectable from 10 weeks of age in *Atp13a2* KO mice, which is substantially earlier than the previously reported motor deficits^2^.

Early onset, widespread gliosis is a key hallmark of Atp13a2 dysfunction in the *Atp13a2* KO mice ^2^. Although unchanged at PND14 (Supplementary Fig. 1e-k), *Atp13a2* KO mice exhibited increased GFAP- and Iba-1-positive areas across multiple brain regions at 10 weeks of age, including the cortex, cerebellum, and striatum (Fig. 1j-k, Supplementary Fig. 1l-q), consistent with early astrogliosis and microgliosis. The area positive for CD68 was additionally increased in the cortex and cerebellum of the KO brain (Fig. 1l-m, Supplementary Fig. 1r), suggesting the presence of more activated microglia. To investigate potential inflammation contributing to this gliosis, we assessed NF-κB signalling in the KO brain as one potential inflammatory pathway ^48^. The mean intensity of nuclear NF-κB was increased in the striatum and cortex of *Atp13a2* KO mice (Supplementary Fig. 1s,t), consistent with increased nuclear translocation. We then asked whether NF-κB nuclear translocation was correlated with altered cytokine levels in CSF of *Atp13a2* KO mice (Fig. 1n). In accordance, levels of pro-inflammatory cytokines including TNF-α and IL-6 were increased, and the anti-inflammatory IL-10 was reduced in the CSF of KO mice (Fig. 1n), indicative of an inflammatory status. Together, these findings support the presence of early brain inflammation upon Atp13a2 dysfunction.

Given the prominent microgliosis, and increased pro-inflammatory cytokine profile in the *Atp13a2* KO mice, we next investigated whether microglia may contribute to this inflammation. Microglia are a major source of pro-inflammatory cytokines including TNF-α and IL-6, largely through NF-κB-dependent signalling^49,50^. Consistent with this role, single cell transcriptomics across neurodegeneration models have revealed conserved microglia states such as disease-associated microglia (DAM) defined by the upregulation of specific genes including *lgals3* and *trem2* ^50–52^. Here, we examined the expression of a focused subset of these neurodegeneration-linked genes in MACS-isolated microglia from *Atp13a2* KO brains (Fig. 1o). Expression of interferon-linked and pro-inflammatory cytokine genes (*Cxcl10, Ifnβ, Il1β*) along with DAM-associated modulators (*lgals3, trem2, apoe, tyrobp, cd68*) was elevated in microglia from *Atp13a2* KO brains relative to WT controls (Fig. 1o). This targeted transcriptional profile suggests that microgliosis in the *Atp13a2* KO brain is associated with transcriptional upregulation of type I interferon and pro-inflammatory linked responses in microglia, which are implicated in neuroinflammation, and neurodegenerative conditions^50–52^.

ATP13A2 loss of function leads to lysosomal polyamine accumulation resulting in the enlargement and accumulation of dysfunctional lysosomes^9,14,15,17^. We assessed whether lysosomal dysfunction resulting from Atp13a2 loss may be correlating with the microglial transcriptional changes and cytokine profile reported in the *Atp13a2* KO brain. The volume and number of Lamp1 positive compartments within microglia was increased in the 10-week-old *Atp13a2* KO striatum (Fig. 1p-r), indicating lysosomal enlargement and dysfunction earlier than previously reported ^2^. *Lgals3*, one of the upregulated genes in *Atp13a2* KO microglia (Fig. 1o), encodes Galectin-3 (Gal-3), a sensor for lysosomal membrane damage mainly expressed and released by reactive microglia in models of neurodegeneration^52–54^. Gal-3 can bind Lamp1 and subsequently accumulate on damaged lysosomes interfering with their clearance, driving inflammation through NF-κB and inflammasome-mediated pathways and downstream cytokine release including TNF-α and IL-6^24^. In this context and in line with lysosomal dysfunction, the volume of Gal-3 co-localised to Lamp1-positive compartments in *Atp13a2* KO microglia was increased compared to WT microglia (Fig. 1s). These findings suggest microglia lysosomal dysfunction may be a potential contributor to the inflammatory state detected in Atp13a2 deficiency.

ATP13A2 is a lysosomal polyamine transporter implicated in the regulation of intracellular polyamine homeostasis^10^. To determine whether polyamine dysregulation precedes behavioural abnormalities, we quantified polyamines in brain, plasma, and liver at PND14, 10 weeks, and 18 months of age using targeted metabolomics^10^ (Fig. 1t-v, Supplementary Fig. 1u-w). At PND14, Put, Spd, and Spm levels were significantly reduced in the brains of *Atp13a2* KO mice (Fig. 1t). These reductions were no longer evident at 10 weeks, whereas by 18 months, Spd, Spm, and N¹-AcSpm levels were increased in KO brains (Fig. 1u,v). In plasma, KO mice exhibited reduced Orn, Spm, and N¹/N⁸-AcSpd levels at PND14, whereas only Spm remained significantly decreased at 10 weeks (Supplementary Fig. 1u,v). In contrast, liver polyamine levels were unchanged at PND14 (Supplementary Fig. 1w), consistent with a more important role for *Atp13a2* in brain polyamine regulation, in line with highest ATP13A2 enrichment in human and mouse brain (Supplementary Fig. 1x,y)

Collectively, these data show that *Atp13a2* deficiency is associated with early-onset motor and non-motor dysfunction, early neuroinflammatory status and lysosomal abnormalities which are preceded by a marked, transient reduction in brain polyamine levels..

### *Atp13a2* deficiency reduces tolerance to polyamine modulation

To assess whether altered polyamine homeostasis contributes to the *Atp13a2* KO phenotype, we examined the effects of negative and positive polyamine modulation on health, motor function, and survival of KO and WT littermates.

To reduce polyamine availability, mice were treated with DFMO, an inhibitor of polyamine synthesis, or with DenSpm, an inducer of polyamine catabolism (Fig. 2a) ^55^. Treatments were initiated at PND2 and continued until 8 weeks of age via *i.p.* injections during early postnatal development and through the drinking water after weaning (Fig. 2a). Neither treatment affected body weight in either genotype, and DFMO did not alter water intake (Supplementary Fig. 2a,b). Notably, DFMO, an FDA-approved drug, is well tolerated in WT mice, but significantly reduced survival of *Atp13a2* KO mice (Fig. 2b). Both DFMO and DenSpm treatments led to earlier onset and more severe motor dysfunction in KO mice, including worsened limb clasping, increased immobility, and enhanced freezing behaviour (Fig. 2c,d and Supplementary Fig. 2c,d).

**Figure 2.**
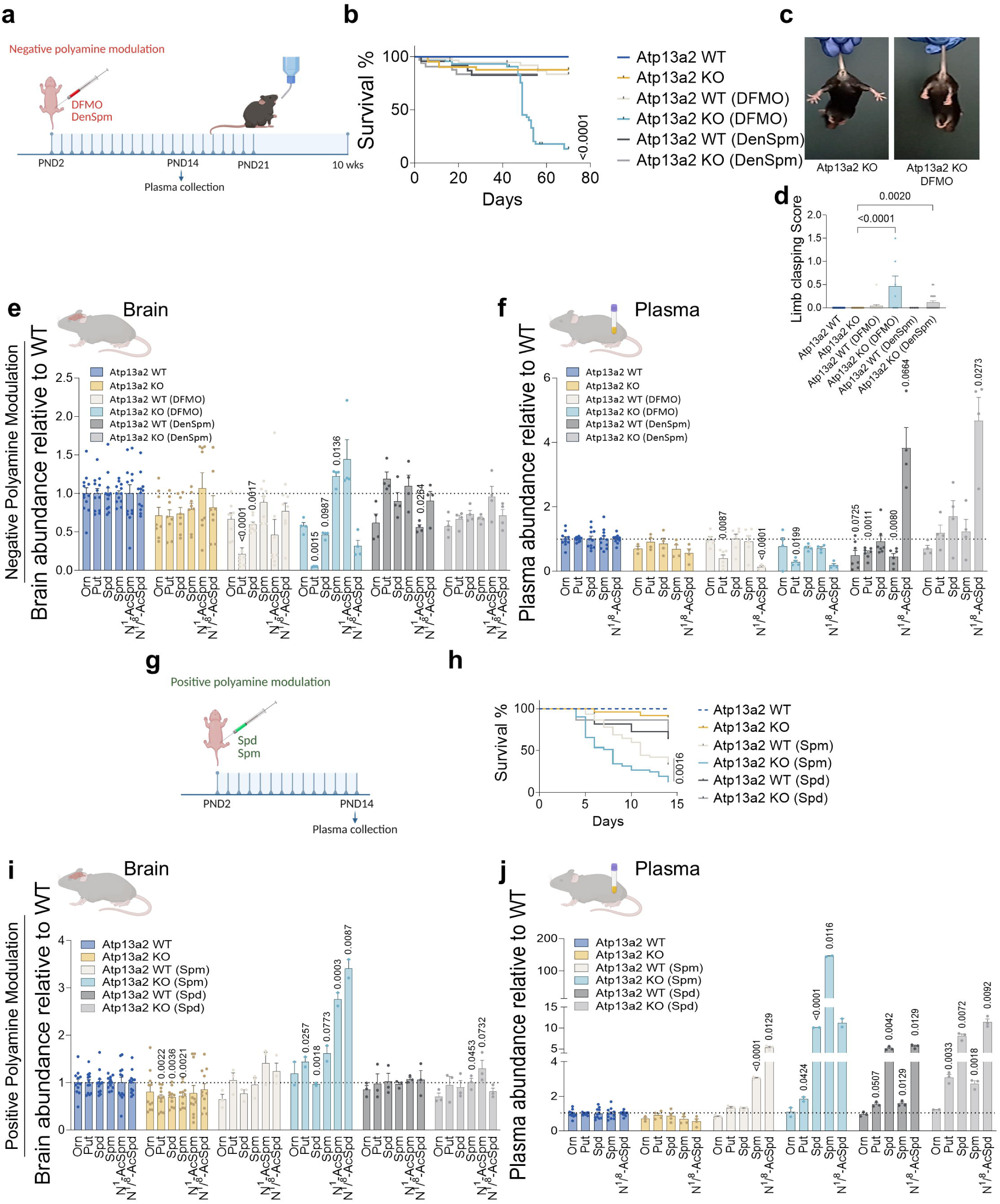
*Atp13a2* deficiency sensitises mice to polyamine modulation. (a) Negative polyamine modulation strategy using DFMO and DenSpm in *Atp13a2* WT and KO mice. (b) Survival of *Atp13a2* WT and KO mice treated with negative polyamine modulators, DFMO and DenSpm. N = 24 WT, 25 WT (DFMO), 5 WT (DenSpm), 28 KO mice, 7 KO (DFMO) and 20 KO (DenSpm). (c) DFMO affects the limb clasping score of *Atp13a2* KO mice. (d) DFMO and DenSpm effects on limb clasping behaviour of *Atp13a2* WT and KO mice. (e) Abundance of polyamine species in the brain of *Atp13a2* WT and KO mice treated with DFMO and DenSpm. N = 12 WT, 8 WT (DFMO), 4 WT (DenSpm), 8 KO mice, 4 KO (DFMO) and 4 KO (DenSpm). (f) Abundance of polyamine levels in the plasma of *Atp13a2* WT and KO mice treated with DFMO and DenSpm. N = 11 WT, 6 WT (DFMO), 7 WT (DenSpm), 4 KO mice, 4 KO (DFMO) and 4 KO (DenSpm). (g) Positive polyamine modulation strategy using Spd and Spm in *Atp13a2* WT and KO mice. (h) Survival of *Atp13a2* WT and KO mice treated with positive polyamine modulators, Spd and Spm. N = 11 WT, 6 WT (Spm), 7 WT (Spd), 4 KO mice, 4 KO (Spm) and 4 KO (Spd). (i) Abundance of polyamine species in the brain of *Atp13a2* WT and *Atp13a2* KO mice treated with Spm and Spd. N = 12 WT, 3 WT (Spm), 3 WT (Spd), 11 KO mice, 2 KO (Spm) and 3 KO (Spd). (j) Abundance of polyamine species in plasma of *Atp13a2* WT and *Atp13a2* KO mice treated with Spm and Spd. N = 11 WT, 3 WT (Spm), 3 WT (Spd), 4 KO mice, 2 KO (Spm) and 3 KO (Spd). Data are represented as mean ± SEM. Log-rank test (b,h). Mixed effects analysis with Tukey’s multiple comparisons post-test (d, e-f, i-j). Ornithine, Orn; Putrescine, Put; Spermidine, Spd; Spermine, Spm; N^1^-Acetylated Spermine, N^1^-AcSpm; N^1^/N^8^-Acetylated Spermidine, N^1^/N^8^-AcSpd. Schematics were created with Biorender.com.

Targeted metabolomics confirmed the reduction of polyamine levels, specifically Put and Spd, by DFMO and DenSpm in plasma and brain at PND14 (Fig. 2e,f). DenSpm increased N¹/N⁸-AcSpd levels in the plasma, consistent with enhanced polyamine catabolism, whereas DFMO reduced N¹/N⁸-AcSpd, reflecting compensatory interplay between synthesis and catabolic pathways^56^. These data indicate that reduction of polyamine availability early in life exacerbates motor impairments and reduces survival in *Atp13a2* KO mice.

To increase polyamine availability, KO and WT mice received *i.p.* injections of Spd or Spm from PND2 to PND14 (Fig. 2g). Spd treatment did not affect survival in either genotype (Fig. 2h), whereas Spm injection reduced survival in WT, in line with earlier reports, although Spm exerts a more pronounced toxicity in KO animals (Fig. 2h)^57,58^.

In line with previous reports^56^, polyamine administration did not significantly alter brain polyamine levels in WT mice (Fig. 2i). In contrast, Spd treatment restored brain polyamine levels in KO mice, whereas Spm treatment resulted in excess brain polyamine accumulation (Fig. 2i). Polyamine injections increased plasma polyamine levels in both genotypes, with a greater increase in Spm and Spd levels observed in KO mice, accompanied by elevated acetylated polyamines, indicative of enhanced degradation (Fig. 2j).

Together, these findings demonstrate that *Atp13a2* loss sensitises mice to both positive and negative polyamine modulation, indicating impaired control of brain and plasma polyamine homeostasis. The early reduction in brain polyamines in KO mice is consistent with a contributory role in behavioural phenotypes.

### Spd supplementation rescues motor and non-motor deficits in *Atp13a2* KO mice

Based on our findings that early-in-life reduction of polyamine levels exacerbates motor dysfunction in *Atp13a2* KO mice, but Spd *i.p.* injection is well tolerated, we next tested whether oral long-term Spd supplementation may ameliorate disease-associated phenotypes. *Atp13a2* KO and WT mice were treated with 3 mM Spd, a concentration previously documented to have beneficial effects against neurotoxicity and neuroinflammation^30,31^, from PND2 until 10 weeks of age, after which motor and non-motor behaviour was assessed at 10 weeks, 6 months, and 12 months (Fig. 3a). Spd treatment did not affect survival in either genotype (Fig. 3b), nor did it alter body weight, food intake, or water consumption (Supplementary Fig. 2e-g), indicating good tolerability. Notably, long-term Spd supplementation robustly rescues motor and non-motor deficits associated with *Atp13a2* deficiency across ages. Spd-treated *Atp13a2* KO mice travelled a total distance in the open field comparable to WTs (Fig. 3c,d). Immobility was restored to WT levels (Fig. 3e), and freezing behaviour was rescued (Supplementary Fig. 2h). In addition, Spd treatment improved average locomotor speed in 12-month-old KO mice (Supplementary Fig. 2i). Spd-treated KO mice also spent significantly more time in the centre of the open field at 10 weeks, 6 months, and 12 months (Fig. 3f), and 10-week-old female KO mice receiving Spd showed a reduced latency to locate the buried pellet (Fig. 3g-i). Importantly, the Spd treatment increased polyamine availability in plasma and brain during early development effectively restoring the polyamine deficit in the *Atp13a2* KO brain at PND14 (Fig. 3j,k). We observed significantly higher Put and Spd levels in *Atp13a2* KO mouse plasma and brain (Fig. 3j,k). Together, our findings demonstrate that oral Spd supplementation impacts plasma and brain polyamine availability and restores motor and non-motor function in *Atp13a2* KO mice.

**Figure 3.**
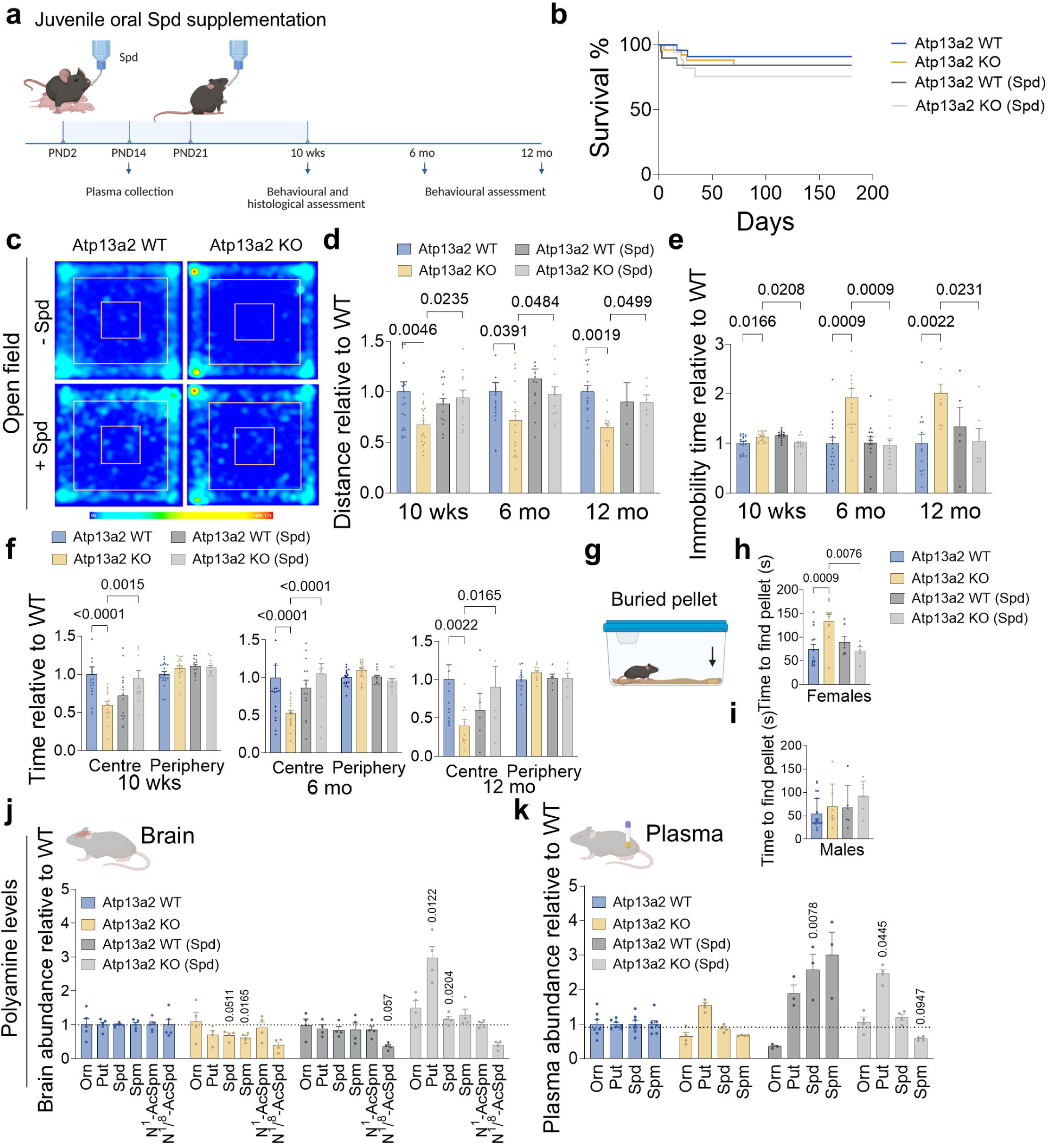
Spermidine supplementation ameliorates motor and non-motor deficits in *Atp13a2* KO mice. (a) Spermidine (Spd) supplementation in *Atp13a2* WT and KO mice from PND2 to 10 weeks of age. (b) Survival of *Atp13a2* WT and KO mice supplemented with Spd or control. N = 24 WT, 28 KO, 19 WT (Spd) and 17 KO (Spd). (c) Representative heatmaps depicting open field areas preferred by *Atp13a2* WT and KO mice with and without Spd supplementation. (d) Total distance traversed by *Atp13a2* WT and KO at 10 weeks, 6 and 12 months. 10 weeks, N = 19 WT, 17 KO, 14 WT (Spd) and 12 KO (Spd). 6 months, N = 19 WT, 17 KO, 13 WT (Spd) and 14 KO (Spd). 12 months, N = 13 WT, 12 KO, 5 WT (Spd) and 6 KO (Spd). (e) Immobility time in the open field. 10 weeks, N = 19 WT, 17 KO, 14 WT (Spd) and 12 KO (Spd). 6 months, N = 19 WT, 17 KO, 13 WT (Spd) and 12 KO (Spd). 12 months, N = 13 WT, 11 KO, 5 WT (Spd) and 5 KO (Spd). (f) Time spent in the centre and periphery of the open field. 10 weeks, N = 19 WT, 17 KO, 19 WT (Spd) and 12 KO (Spd). 6 months, N = 16 WT, 17 KO, 12 WT (Spd) and 12 KO (Spd). 12 months, N = 13 WT, 20 KO, 5 WT (Spd) and 8 KO (Spd). (g) Illustration depicting the buried pellet test. (h) Time taken for female mice to locate the pellet in the buried pellet test at 10 weeks of age. N = 10 WT, 13 KO, 7 WT (Spd) and 5 KO (Spd). (i) Time taken by male mice at 10 weeks of age to locate the hidden pellet. N = 11 WT, 7 KO, 6 WT (Spd) and 7 KO (Spd). (j) Abundance of polyamine species in the brain at PND14. N = 5 WT, 4 KO, 4 WT (Spd) and 4 KO (Spd). (k) Abundance of polyamine species in the plasma at PND14. N = 7 WT, 4 KO, 3 WT (Spd) and 4 KO (Spd). Data are represented as mean ± SEM. Log-rank test (b). Two-way ANOVA with Bonferroni multiple comparisons post-test (d-f). One-way ANOVA with Tukey’s post-test (h-i). Mixed effects analysis with Tukey’s post-test (j-k). Ornithine, Orn; Putrescine, Put; Spermidine, Spd; Spermine, Spm; N^1^-Acetylated Spermine, N^1^-AcSpm; N^1^/N^8^-Acetylated Spermidine, N^1^/N^8^-AcSpd. Illustrations were made using Biorender.com.

To determine whether these effects were specific to Spd, parallel experiments were performed with oral Spm supplementation. Spm treatment did not affect body weight, food, water intake, or survival (Supplementary Fig. 2e-g and Supplementary Fig. 3a,b). However, Spm failed to improve motor performance in *Atp13a2* KO mice across all ages as measured in the total distance travelled, immobility, freezing behaviour, and average speed (Supplementary Fig. 3c-g). Spm supplementation was also unable to rescue non-motor deficits, including the reduced time in the centre of the open field or impaired buried pellet retrieval (Supplementary Fig. 3h-k). Despite this lack of behavioural improvement, Spm treatment increased plasma and brain Put and Spd levels in KO mice, but had a stronger effect on Spm accumulation in the brain, which may have offset any beneficial effect (Supplementary Fig. 3l,m).

Collectively, these data demonstrate that Spd, but not Spm, restores motor and non-motor function in *Atp13a2* KO mice, supporting a contributory role for early polyamine deficiency in the behavioural phenotype.

### Juvenile Spd supplementation rescues neuroinflammation in *Atp13a2* KO mice

We next examined whether oral juvenile Spd supplementation may modulate the early reactive gliosis, transcriptional and lysosomal changes in *Atp13a2* KO mice at 10 weeks of age, a time point at which motor and non-motor deficits are fully rescued (Fig. 3).

Juvenile supplementation of Spd (Fig. 4a-g), but not Spm (Supplementary Fig. 4a-o), prevented the reported increase in GFAP, Iba-1, CD68 positive area across multiple regions of the *Atp13a2* KO brain, when assessed against the same control cohort in a single experiment. Spd reduced GFAP-positive area in the cortex, cerebellum and striatum to levels comparable to those observed in *Atp13a2* WT mice (Supplementary Fig. 4a-d). Similarly, Iba-1-positive (Fig. 4a-d) and CD68-positive (Fig. 4e-g) areas were significantly decreased in Spd-treated *Atp13a2* KO brains. Notably, Spd treatment also reduced Iba-1-positive area in *Atp13a2* WT mice relative to untreated controls (Fig. 4b-d), suggesting a strong modulatory effect on microglia. Juvenile Spd supplementation also reduced NF-κB nuclear translocation in the brains of KO mice (Supplementary Fig. 4p-r), and fully normalised the expression of interferon-linked and pro-inflammatory cytokine genes (*Cxcl10, Ifnβ, Il1β*) along with DAM-associated modulators (*lgals3, trem2, tyrobp, cd68*) elevated in *Atp13a2* KO microglia (Fig 4h-p).

**Figure 4.**
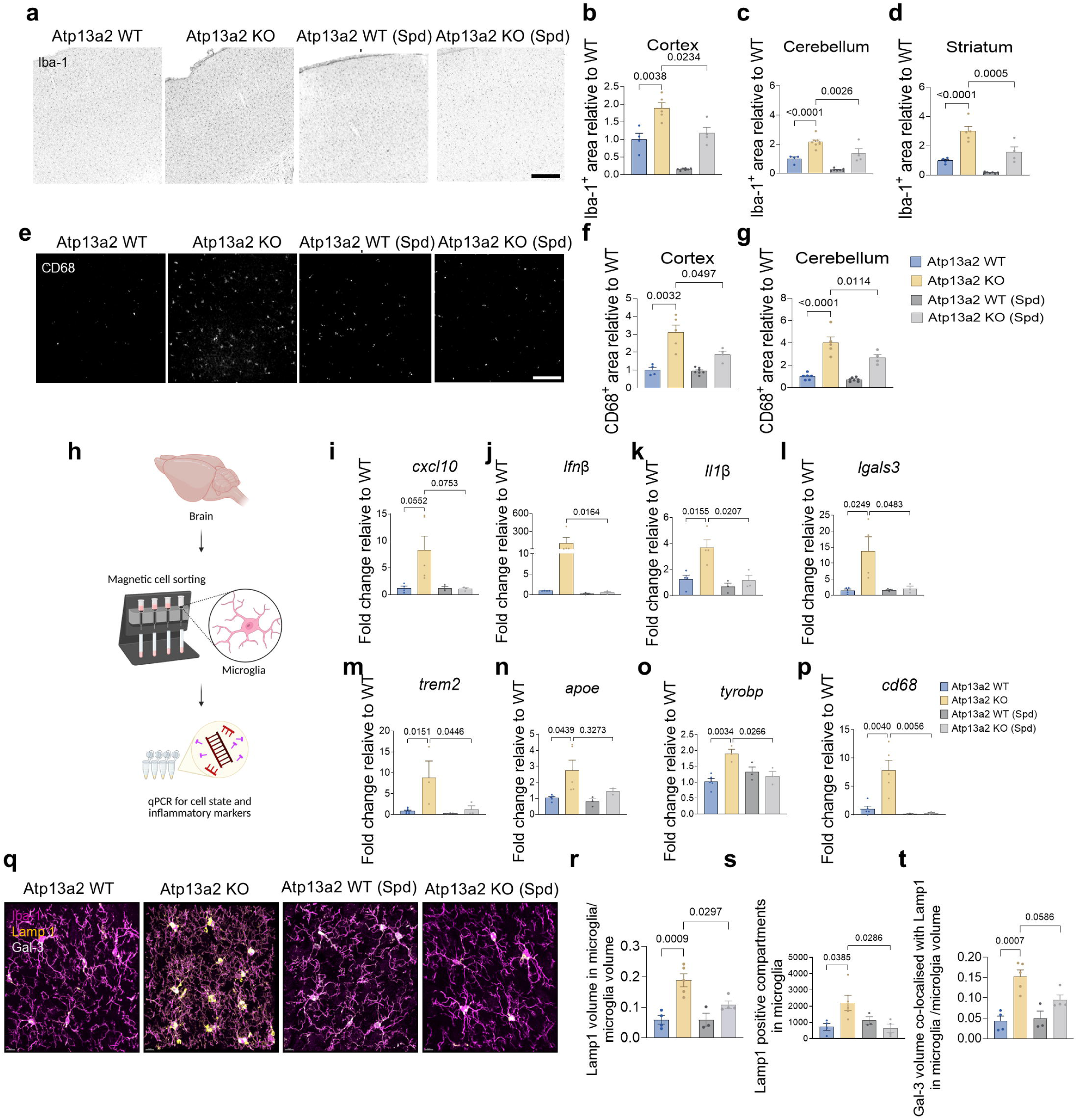
Juvenile spermidine supplementation rescues neuroinflammation in the brains of *Atp13a2* KO mice. (a) Representative images of cortical microglia (Iba-1, black) of *Atp13a2* WT and KO mice given spermidine (Spd) or vehicle control. Scale bar = 200 µm. (b) The area positive for Iba-1 in the cortex. N = 4 WT, 5 KO, 7 WT (Spd) and 4 KO (Spd). (c) Iba-1 positive area in the cerebellum. (d) The area positive for Iba-1 in the striatum. N = 4 WT, 5 KO, 7 WT (Spd) and 4 KO (Spd). (e) Representative images of cortical microglia (CD68, white) of *Atp13a2* WT and KO mice treated with Spd. Scale bar = 200 µm. (f) The area positive for CD68 in the cortex. N = 4 WT, 5 KO, 7 WT (Spd) and 4 KO (Spd). (g) CD68 positive area in the cerebellum. N = 5 WT, 5 KO, 7 WT (Spd) and 4 KO (Spd). (h) Isolation and assessment of gene expression of microglia from 10-weeks-old *Atp13a2* WT and KO mice treated with Spd or vehicle control. (i-p) Effect of Spd supplementation on the expression of a subset of pro-inflammatory and disease-associated genes in *Atp13a2* WT and KO microglia. N = 4-5 WT, 4 KO, 3 WT (Spd) and 3 KO (Spd). (q) Representative images of Lamp1 (yellow) and Galectin-3 (Gal-3, white) within striatal microglia (Iba-1, magenta) of *Atp13a2* WT and KO mice treated with Spd or vehicle control. Scale bar = 10 µm. (r) Lamp1 volume per microglia volume of *Atp13a2* WT and KO striatum. N = 4 WT, 5 KO, 3 WT (Spd) and 4 KO (Spd). (s) Number of Lamp1 positive compartments in striatal microglia of *Atp13a2* WT and KO mice. N = 4 WT, 5 KO, 3 WT (Spd) and 4 KO (Spd). (t) Volume of Gal-3 co-localised with Lamp1 per microglia volume. N = 4 WT, 5 KO, 3 WT (Spd) and 4 KO (Spd). Data are represented as mean ± SEM. One-way ANOVA with Šídák’s (b-d, f-g), Tukey’s (i,l-o,r-t) and Bonferroni (k,p) multiple comparisons post-tests. Kruskal-Wallis test (j) Illustrations were created on Biorender.com.

Spd positive outcomes on Iba-1 and CD68 positive areas and microglia transcription changes were associated with a reduction in the number and volume of Lamp1 positive compartments and a marked reduction in Gal-3 volume co-localised with Lamp1 in KO microglia in line with improved lysosomal health (Fig. 4q-t). Spm on the contrary failed to restore lysosomal health in microglia, and even increased the volume of Lamp1 positive compartments in the WT striatum and Gal-3 volume in the KO brain (Supplementary Fig. 4s-u).

Together, these data demonstrate that juvenile Spd, but not Spm, supplementation prevents astrogliosis and microgliosis in *Atp13a2* KO mouse brains, which is associated with suppression of pro-inflammatory, interferon-linked microglial responses and enhanced lysosomal health.

### Spd supplementation mediates anti-inflammatory actions through Atp13a3

To investigate the mechanism behind the anti-inflammatory effect of Spd in the context of *Atp13a2* deficiency, we turned to primary microglia of *Atp13a2* KO and WT mice and monitored the morphology as a readout of microglia pro-inflammatory activation (resting conditions, elongated shape/high aspect ratio; inflamed state, rounded shape/low aspect ratio). *Atp13a2* KO microglia presented a lower aspect ratio compared to *Atp13a2* WT counterparts (Fig. 5a-c), and increased TNF-α levels (Fig. 5d) in line with an inflamed state under basal conditions ^40^ pointing to a cell-autonomous mechanism of inflammation. The inflammatory stimulus LPS significantly lowered the aspect ratio in *Atp13a2* WT and KO microglia promoting the inflammatory state. Notably, WT microglia morphology was significantly restored to control levels when exogenous Spd was supplied, illustrating that Spd mediates an anti-inflammatory effect under LPS-treated conditions. A Spd rescue effect was also observed in *Atp13a2* KO microglia, but this required a higher concentration than for WT microglia (Fig. 5b-c). Spd also restored cytokine expression and secretion (Fig. 5d and Supplementary Fig. 5 a-d), demonstrating the potent anti-inflammatory effect of Spd on WT and KO microglia in culture. Since this *in vitro* model recapitulates the protective effects of Spd on brain neuroinflammation *in vivo*, we used it to further study the mechanism of Spd rescue on the neuroinflammatory response.

**Figure 5.**
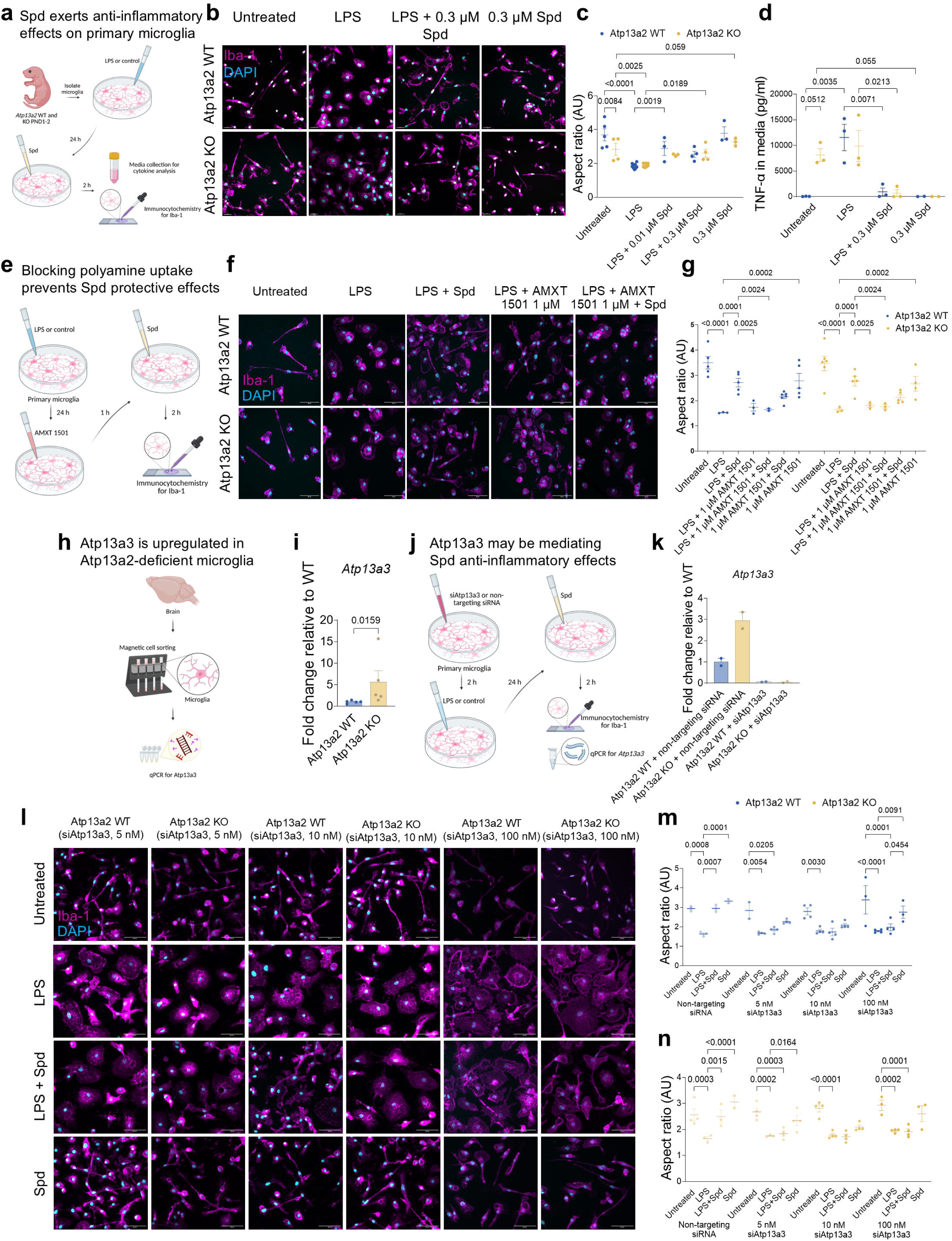
Spermidine mediates anti-inflammatory effects in isolated mouse microglia through Atp13a3. (a) Lipopolysaccharide (LPS) and spermidine (Spd) treatments of cultured primary microglia from *Atp13a2* WT and KO mice. (b) Maximum projection confocal images of *Atp13a2* WT and KO microglia (Iba-1, magenta) treated with LPS and 0.3 µM Spd. DAPI (cyan); Scale bar = 20 µm. (c) Aspect ratio of *Atp13a2* WT and KO microglia exposed to a combination of LPS and Spd (0.01 and 0.3 µM). N = 5 WT, 4 KO, 6 WT (LPS), 6 KO (LPS), 3 WT (0.01 µM + LPS), 3 KO (0.01 µM + LPS), 4 WT (0.3 µM + LPS), 4 KO (0.3 µM + LPS), 4 WT (0.3 µM Spd) and 4 KO (0.3 µM Spd). (d) Concentration of TNF-α secreted in the media of *Atp13a2* WT and KO microglia treated with LPS and 0.3 µM Spd. N = 3 WT, 3 KO, 3 WT (LPS), 3 KO (LPS), 3 WT (0.3 µM + LPS), 3 KO (0.3 µM + LPS), 2 WT (0.3 µM Spd) and 2 KO (0.3 µM Spd). (e) Illustration depicting AMXT 1501 treatment of *Atp13a2* WT and KO microglia. (f) Maximum projections of *Atp13a2* WT and KO microglia (Iba-1, magenta) treated with AMXT 1501 in combination with LPS or Spd. DAPI (cyan); Scale bar = 50 µm. (g) Aspect ratio of *Atp13a2* WT and KO microglia treated with a combination of LPS, AMXT 1501 and Spd. N = 5 WT, 6 KO, 3 WT (LPS), 3 KO (LPS), 6 WT (0.3 µM Spd + LPS), 6 KO (0.3 µM Spd + LPS), 5 WT (LPS + 1 µM AMXT 1501), 3 KO (LPS + 1 µM AMXT 1501), 4 WT (LPS + 1 µM AMXT 1501+ 0.3 µM Spd), 4 KO (LPS + 1 µM AMXT 1501 + 0.3 µM Spd), 6 WT (1 µM AMXT 1501 + 0.3 µM Spd), 6 KO (1 µM AMXT 1501 + 0.3 µM Spd), 5 WT (1 µM AMXT 1501) and 4 KO (1 µM AMXT 1501). (h) Illustration of magnetic cell sorting of cortical microglia from 10-week-old *Atp13a2* WT and KO brains. (i) mRNA expression of *Atp13a3* in the 10-weeks-old *Atp13a2* WT and KO isolated microglia. N = 5 WT and 4 KO. (j) Knocking down *Atp13a3* (siAtp13a3) in isolated *Atp13a2* WT and KO microglia and subsequent treatment. (k) Expression of *Atp13a3* in isolated microglia treated with siAtp13a3 (10 nM) or non-targeting siRNA (10 nM). N = 2 WT (non-targeting siRNA), 2 WT (siAtp13a3), and 2 KO (non-targeting siRNA) and 2 KO (siAtp13a3). (l) Maximum projection images of *Atp13a2* WT and KO microglia (Iba-1, magenta) treated with siAtp13a3 (5, 10 or 100 nM) in combination with LPS, or Spd. DAPI (cyan); Scale bar = 20 µm. (m) Aspect ratio of *Atp13a2* WT microglia treated with siAtp13a3 (5, 10 or 100 nM) or non-targeting siRNA (10 nM) in combination with LPS and Spd. N = 3 WT (non-targeting siRNA), 3-4 WT ( 5nM - 100 nMsiAtp13a3). (n) Aspect ratio of *Atp13a2* KO microglia. N = 3 KO (non-targeting siRNA), 3 KO ( 5, 10 and 100 nM siAtp13a3). Data are represented as mean ± SEM. Mixed effects analysis with Tukey’s multiple comparisons post-test (c-d,g,m-n). Mann-Whitney test (i). Schematics were made on Biorender.com.

Mitochondrial oxidative stress is closely coupled to microglia inflammatory activation and may act both upstream and downstream of neuroinflammatory signalling, involving activation of IL1β, CXCL10, IFNβ ^59–62^. Since ATP13A2 counters mitochondrial oxidative stress by transporting polyamines from lysosome to mitochondria offering an antioxidant response ^11^, we examined whether microglial activation and Spd rescue may coincide with changes in mitochondrial oxidative stress. We detected elevated levels of mitochondrial reactive oxidative species (mitoROS; measured via mitoSox) in basal conditions and after LPS treatment in *Atp13a2* KO microglia (Supplementary Fig. 5e-g), which were reduced by exogenous supplementation of Spd, mimicking the effects of the mitochondrial anti-oxidant mitoTEMPO (MT, Supplementary Fig. 5f,g). Our results indicate that *Atp13a2* deficiency causes increased mitochondrial oxidative stress, which is effectively counteracted by Spd or MT supplementation in basal or LPS conditions, suggesting that Spd mitigates microglia inflammatory activation in association with reduced mitochondrial oxidative stress.

A key mechanistic question is whether exogenous Spd can enter cells in the absence of functional Atp13a2 or instead may exert its effects extracellularly. To address this, we inhibited cellular polyamine uptake using AMXT 1501, a broad polyamine transport inhibitor. AMXT 1501 abolished the anti-inflammatory effects of exogenous Spd on microglia morphology, indicating that Spd must be transported into cells via an AMXT 1501-sensitive uptake mechanism (Fig. 5e-g). We recently showed that AMXT 1501 targets ATP13A3, with ATP13A3 representing a major contributor to cellular polyamine uptake^42,63^. We therefore investigated whether Atp13a3 mediates Spd-dependent anti-inflammatory responses in cultured microglia. Expression analysis revealed increased Atp13a3 levels in MACS-isolated and primary microglia from *Atp13a2* KO mice compared with WT controls, suggesting compensatory upregulation in the absence of *Atp13a2* (Fig. 5h,i). When *Atp13a3* was selectively silenced in cultured *Atp13a2* KO and WT microglia, the Spd-induced anti-inflammatory response was prevented in both genotypes, an effect not observed with a non-targeting siRNA control (Fig. 5j-n). These findings demonstrate that Atp13a3-mediated uptake is required for Spd to exert anti-inflammatory effects in microglia.

Finally, we examined whether Spd-mediated modulation of microglia inflammatory state influences neuronal morphology with microglia-neuron co-cultures (Supplementary Fig. 5h-j). Co-culturing WT microglia with WT neurons had no effect on neurite length (Supplementary Fig. 5i,j). On the contrary, co-culture of *Atp13a2* KO microglia with WT neurons significantly reduced neurite length (Supplementary Fig. 5 i,j). In line with Spd protective effects, Spd-treated *Atp13a2* KO microglia did not exert this detrimental effect on neurite length (Supplementary Fig. 5i,j). These results indicate that activated *Atp13a2*-deficient microglia negatively impact neuronal morphology, and that this effect is prevented by Spd treatment.

Together, these data provide a mechanistic link between Spd uptake via Atp13a3, mitochondrial antioxidant response and suppression of the inflammatory state, and neuron-glia interactions that may contribute to the neuroinflammatory and behavioural phenotypes observed *in vivo*.

### Spd-mediated rescue across models of Kufor-Rakeb syndrome and parkinsonism

In line with the prominent astrogliosis and microgliosis in the brain of *Atp13a2* knockout mice, we observed astrogliosis and microgliosis also in post-mortem brain tissue from a PARK-ATP13A2 patient carrying a compound heterozygous pathogenic *ATP13A2* variant (patient II.14; Table 1)^3,27^, whose fibroblasts has been previously shown to exhibit mitochondrial dysfunction^16^. CD68, Iba-1 and GFAP staining was assessed in the frontal cortex (FC), the midbrain including the substantia nigra (SN), and the striatum (Th) (Fig. 6a). Astrogliosis was severe in the central grey and midbrain (Fig. 6a), but less in cortical areas (Fig. 6a). Microglia activation was seen in the frontal cortex, the midbrain including the substantia nigra, and the striatum (Fig. 6a) with reactive CD68-positive microglia pronounced in the central grey and midbrain (Fig. 6a).

**Figure 6.**
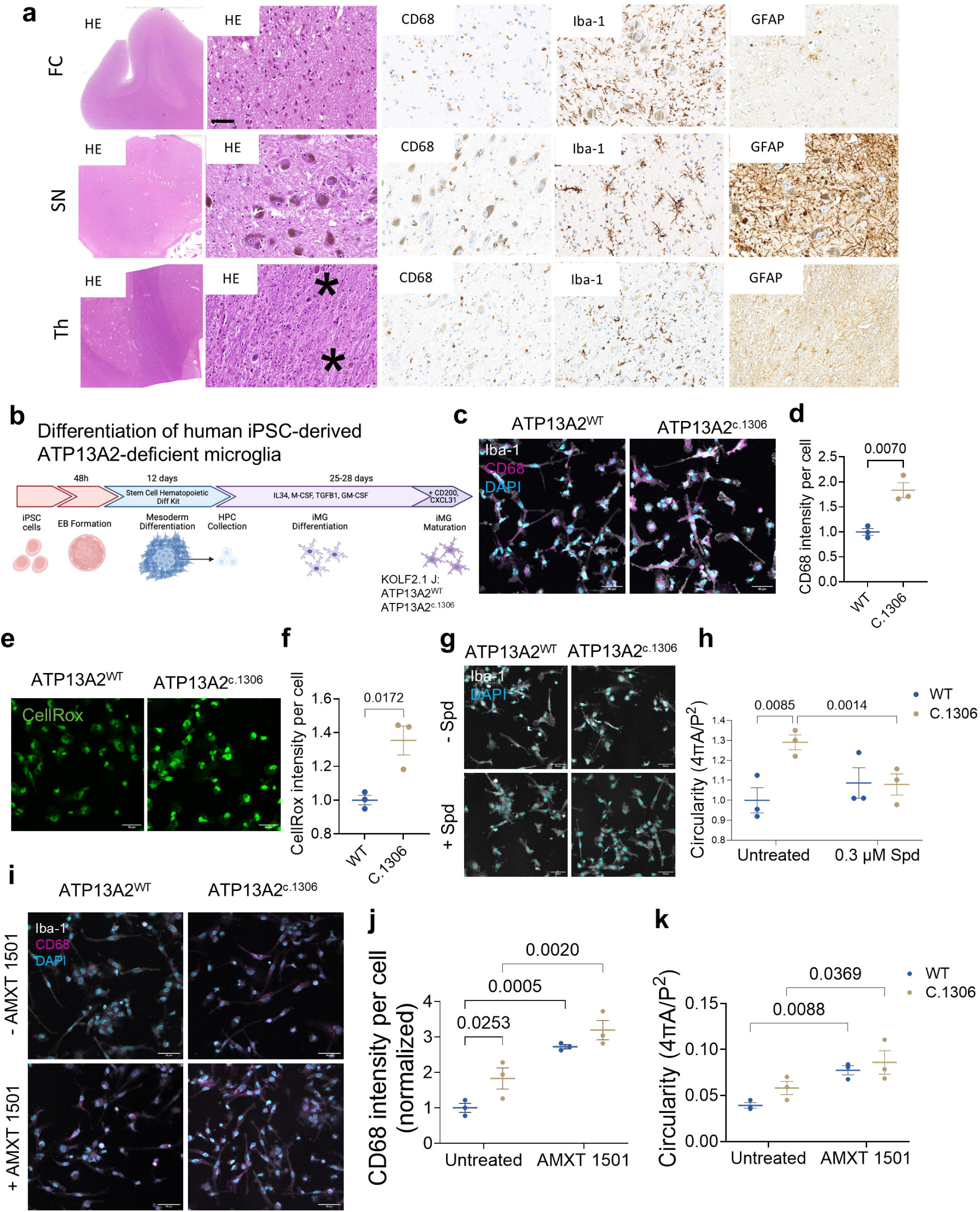
ATP13A2 dysfunction induces gliosis in human postmortem brain and human iPSC-derived microglia. (a) Immunohistochemical analysis of brain tissue from a KRS patient carrying a pathogenic ATP13A2 variant. Haematoxylin and eosin (HE) staining in the frontal cortex (FC), the midbrain including the substantia nigra (SN), and the striatum (Th). CD68, Iba-1 and GFAP immunoreactivity. Scale bar = 50 μm. (b) Differentiation of human iPSC-derived ATP13A2-deficient (ATP13A2^c.1306^) and ATP13A2^WT^ microglia. (c) Representative images of CD68 (magenta) in induced ATP13A2^c.1306^ and ATP13A2^WT^ microglia (Iba-1, gray; DAPI, cyan). Scale bar = 50 µm. (d) CD68 intensity per ATP13A2^c.1306^ and ATP13A2^WT^ microglia. N = 3 biological replicates of ATP13A2^c.1306^ and 3 ATP13A2^WT^. (e) Representative images of CellRox in ATP13A2^c.1306^ and ATP13A2^WT^ microglia. Scale bar = 50 µm. (f) CellRox intensity in ATP13A2^c.1306^ and ATP13A2^WT^ microglia. N = 3 biological replicates of ATP13A2^c.1306^ and 3 ATP13A2^WT^. (g) Representative images of Spd or control treated ATP13A2^c.1306^ and ATP13A2^WT^ iPSC-derived microglia (Iba-1, gray; DAPI, cyan). Scale bar = 50 µm. (h) Circularity of Spd or control treated ATP13A2^c.1306^ and ATP13A2^WT^ microglia. N = 3 biological replicates of ATP13A2^c.1306^ and 3 ATP13A2^WT^. (i) Representative images of CD68 (magenta) in AMXT 1501 treated ATP13A2^c.1306^ and ATP13A2^WT^ microglia (Iba-1, gray; DAPI, cyan). (j) CD68 intensity per ATP13A2^c.1306^ and ATP13A2^WT^ microglia. N = 3 biological replicates of ATP13A2^c.1306^ and 3 ATP13A2^WT^. (k) Circularity of AMXT 1501 treated ATP13A2^c.1306^ and ATP13A2^WT^ microglia. N = 3 biological replicates of ATP13A2^c.1306^ and 3 ATP13A2^WT^. Data are represented as mean ± SEM. Unpaired student’s t-test (d,f). Mixed effects ANOVA (h,j,k)

**Table 1.** Summary of clinical features of the patient II.14 with PARK-ATP13A2, adapted from Behrens *et al.*^27^.

|  |  |
| --- | --- |
| Age of onset (yr) | 10 - 14 |
| Age at death (yr) | 55 - 59 |
| Duration of disease (yr) | 45 - 49 |
| *ATP13A2* pathogenic variants | c.3057delC/ IVS13 5:G A |
| Clinical signs | Slowly progressive juvenile-onset parkinsonism with pyramidal signs, supranuclear gaze palsy, cognitive decline, mini-myoclonus and dystonia |

To assess the translational relevance of our findings with Spd treatment in *Atp13a2*-deficient mice, we turned to human iPSC-derived microglia harbouring the ATP13A2 c.1306 loss-of-function frameshift mutation (ATP13A2^c1306^) that is associated with KRS and that results in a functional knockout of ATP13A2 ^64^. Induced ATP13A2^c1306^ microglia showed increased CD68 expression (Fig. 6b-d), cellular reactive oxidative species (CellRox; Fig. 6e,f), and increased circularity (Fig. 6g,h), suggestive of an inflammatory state under basal conditions. Exogenous Spd supplementation significantly lowered the circularity (Fig. 6g,h), in line with an anti-inflammatory effect of Spd on iPSC-derived ATP13A2^c1306^ microglia. Consistent with a role of exogenous polyamine dysregulation in neuroinflammation onsets, AMXT 1501 exacerbated the circularity and CD68 phenotypes in ATP13A2^c1306^ microglia, and an inflammatory-like state in ATP13A2^wt^ (Fig. 6i-k). These findings provide further evidence for a cell-autonomous mechanism underlying neuroinflammation in ATP13A2-deficiency, and support the translational relevance of Spd supplementation in a human context.

To explore whether Spd supplementation may have therapeutic potential beyond early disease stages, we next assessed the effects of Spd treatment initiated in adulthood. *Atp13a2* KO mice were treated with Spd from 7 to 12 months of age, followed by behavioural assessment (Fig. 7a). Adult-onset Spd supplementation significantly increased total distance travelled in the open field and reduced immobility and freezing behaviour in *Atp13a2* KO mice (Fig. 7c-e). Time spent in the centre of the open field was also increased in both KO and WT mice receiving Spd (Fig. 7f). These findings indicate that Spd supplementation can ameliorate motor and non-motor phenotypes associated with *Atp13a2* deficiency even when initiated at later stages of disease progression.

**Figure 7.**
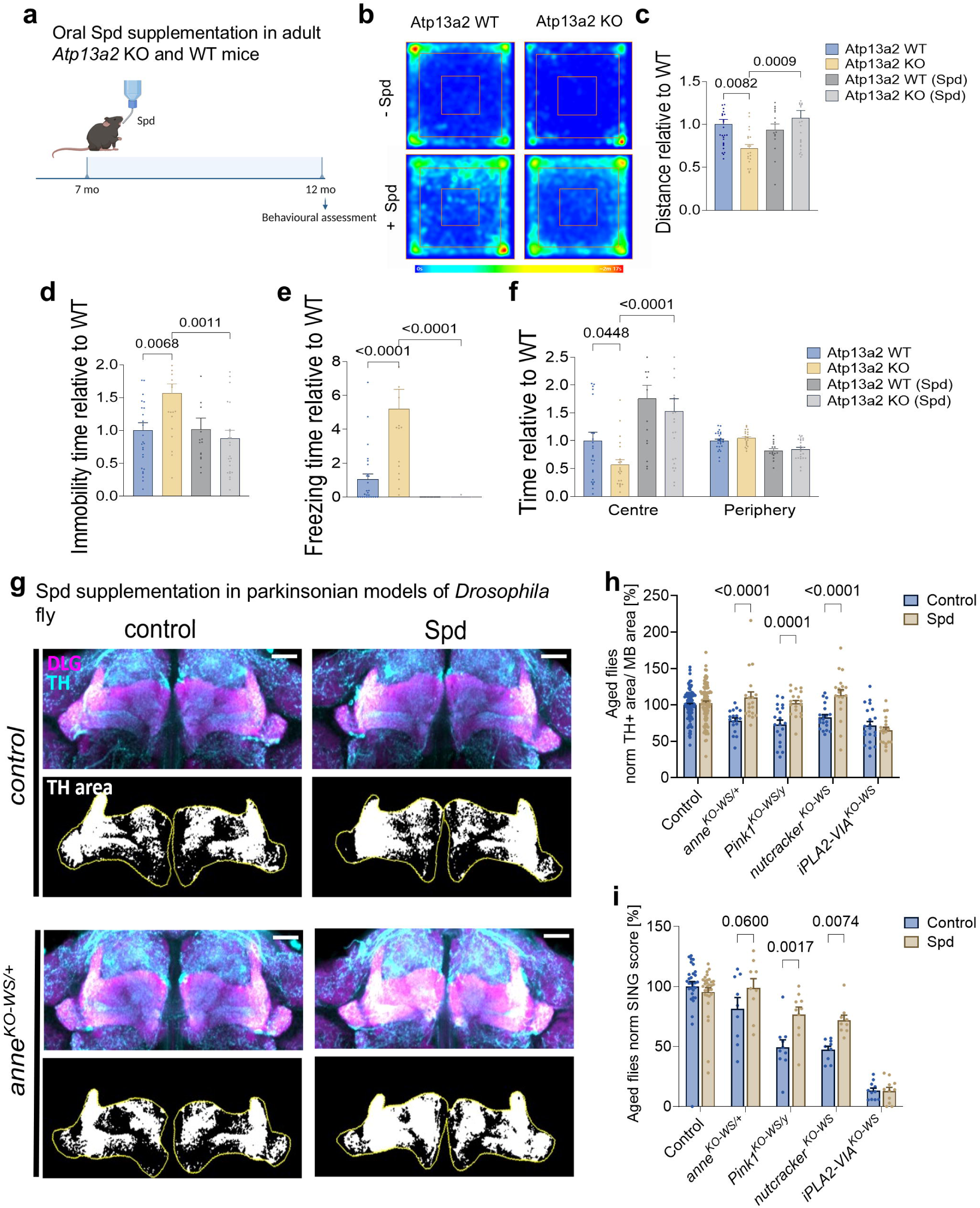
Spermidine-mediated rescue across models of Kufor-Rakeb syndrome and parkinsonism. (a) Timeline for spermidine (Spd) supplementation in adult *Atp13a2* WT and KO mice. (b) Heatmaps depicting the preferred zones of *Atp13a2* WT and KO mice in the open field at 12 months. (c) Total distance traversed by *Atp13a2* WT and KO mice. N = 27 WT, 20 KO, 15 WT (Spd) and 24 KO (Spd). (d) Immobility time. N = 27 WT, 20 KO, 15 WT (Spd) and 24 KO (Spd). (e) Freezing time. N = 27 WT, 20 KO, 15 WT (Spd) and 24 KO (Spd). (f) Time in the centre and periphery. N = 27 WT, 20 KO, 15 WT (Spd) and 24 KO (Spd). (g) Maximum projection confocal images of aged control (*CS^w1118^w+*)and *anne^KO-WS/+^* fly brains treated with 5 mM Spd or control visualising dopaminergic innervation (anti-TH, cyan) and the post-synaptic zone of mushroom body, MB (anti-DLG, magenta) and TH+ area of “middle z-plane” within the MB (yellow, outline of MB). Scale bar = 20 µm. (h) Quantification of dopaminergic synaptic area within the MB area in aged fly models treated with Spd or solvent-control normalised (norm) to the control. (i) SING score for aged fly models treated with Spd or control. Data are represented as mean ± SEM. One-way ANOVA with Šídák’s (c), Tukey’s (d) and Dunnett’s (e) multiple comparisons tests. Two-way ANOVA with Dunnett’s (f) and Tukey’s (h,i) multiple comparisons tests. Illustrations were generated on Biorender.com.

To assess the broader translational relevance of Spd treatment in parkinsonian disorders associated with dopaminergic neuron loss, we next examined *Drosophila melanogaster* loss-of-function models of early-onset parkinsonism, including heterozygous *anne^KO-WS/+^* (*Atp13a2* fly ortholog, *PARK9),* hemizygous null *Pink1^KO-WS^* (*PARK6*), *nutcracker^KO-WS^* (*FBXO7*), and homozygous KO *iPLA2-VIA^KO-WS^* (*PARK14*). These models exhibit age-dependent motor decline in the SING assay, associated with dopaminergic dysfunction and reduced dopaminergic innervation of the MB^45^. In aged flies, all four models displayed significantly reduced tyrosine hydroxylase (TH)-positive dopaminergic innervation of the MB compared to *CS^w1118^w^+^* control flies (Fig. 7g,h). Notably, Spd treatment restored dopaminergic innervation in *anne^KO-WS/+^, nutcracker^KO-WS^,* and *Pink1^KO-WS^* flies, which was accompanied by partial improvement in SING performance (Fig. 7h,i). In contrast, Spd treatment did not rescue dopaminergic deficits or motor performance in *iPLA2-VIA^KO-WS^* flies (Fig. 7h,i). These data suggest that Spd selectively ameliorates dopaminergic neuron dysfunction or loss in a subset of genetic parkinsonism models linked to ATP13A2, FBXO7 and PINK1 deficiency.

Together, our findings demonstrate that Spd rescues disease-associated phenotypes in multiple models of ATP13A2 deficiency and is associated with improved neuronal (including dopaminergic neuron) integrity, highlighting the translational potential of polyamine modulation in Kufor-Rakeb syndrome and possibly a subset of related parkinsonian disorders.

## Discussion

Our study identifies early brain polyamine deficiency as a mechanistic link between ATP13A2 loss, lysosomal damage, mitochondrial oxidative stress, neuroinflammation, and (non-)motor dysfunction, which can be reversed by Spd supplementation across model systems providing new therapeutic angles for Kufor-Rakeb syndrome and possibly other parkinsonian subtypes. Our work expands the scope of ATP13A2 pathology, redefining it as a glia-mediated metabolic inflammatory disorder in addition to neuronal lysosomal storage disease.

### Neuroinflammation and glial-mediated neuronal vulnerability in ATP13A2 pathology

In this study, we report glial dysfunction as a key contributor to neuronal vulnerability in ATP13A2-associated pathology. Astrogliosis and microgliosis are consistent features of ATP13A2 deficiency across species and model systems, including Atp13a2-deficient mice, dogs, rats and non-human primate models^2,20–22^. We describe for the first time reactive gliosis in a KRS patient carrying a loss-of-function ATP13A2 mutation. Notably, glial activation arises early and precedes motor and non-motor phenotypes in *Atp13a2*-deficient mice, supporting a role in disease initiation rather than a secondary response to neurodegeneration.

At the cellular level, ATP13A2-deficient microglia adopt an activated phenotype characterized by NF-κB signaling, increased pro-inflammatory cytokine production, and induction of a disease-associated-microglia(DAM)-like gene program. While DAM signatures may initially reflect adaptive responses, our data suggest that chronic ATP13A2 deficiency shifts microglia state towards a maladaptive, neurotoxic phenotype as a consequence of lysosomal and mitochondrial stress. Specifically, Atp13a2 deficiency induces lysosomal dysfunction and mitochondrial oxidative stress in mouse microglia, consistent with previous findings^10,11,17^, which together lower the threshold for inflammatory activation ^24,26,65^ marked by activation of NF-κB signaling and enhanced pro-inflammatory cytokine production. In a parallel study, NLRP3 inflammasome activation in ATP13A2-deficient macrophages and microglia has independently been mechanistically connected to lysosomal and mitochondrial dysfunction^66^.

Beyond these cell-autonomous effects, our co-culture experiments demonstrate that Atp13a2-deficient microglia actively promote neuronal vulnerability, impairing neuronal morphology and reducing neurite length. Given that microglia are key regulators of neuronal circuitry^23,67^, and that neurite retraction represents an early hallmark of neurodegeneration^68–70^, our results position microglia as active contributors to early neuronal dysfunction.

Astrocytes likely reinforce this process, since prominent astrogliosis is observed in *Atp13a2* KO mice. Murine *Atp13a2*-deficient astrocytes exhibit NLRP3 inflammasome activation as a consequence of lysosomal membrane permeabilization^65^, and in a parallel study we demonstrate that human ATP13A2-deficient iPSC-derived astrocytes acquire a reactive phenotype and secrete toxic factors detrimental to neurons^64^. Together, these findings support a disease mechanism in which ATP13A2 loss engages an inflammatory program, in which both microglia and astrocytes converge to amplify neuronal vulnerability. This framework may explain why early motor and non-motor symptoms emerge following the onset of neuroinflammation.

Overall, our mechanistic insights provide a unifying framework linking lysosomal dysfunction, mitochondrial oxidative stress, glial activation, and neuronal impairment in ATP13A2-associated pathology.

### Disrupted lysosomal polyamine flux drives neuroinflammation via polyamine depletion

Loss-of-function variants in the endo-lysosomal polyamine transporter ATP13A2 cause Kufor-Rakeb syndrome, a juvenile-onset parkinsonism manifesting as early as 12 years of age ^7^. However, how impaired polyamine transport contributes to early disease onset has remained unclear. In cells, ATP13A2 functions as a lysosomal polyamine exporter that contributes to cellular polyamine uptake^10^. Here, we identify an early and transient reduction in brain polyamine levels as a previously unrecognized pathogenic feature of ATP13A2 deficiency that precedes neuroinflammation and the onset of motor and non-motor abnormalities. At later stages, total brain polyamine levels gradually increase, consistent with parallel reports ^17,71^. However, our *in vivo* polyamine modulation experiments clearly demonstrate that reduced polyamine availability is a key driver of disease onset and progression. Pharmacological depletion of polyamines exacerbates motor dysfunction and reduces survival in *Atp13a2* knockout mice, indicating a critical dependence on cytosolic polyamine availability when lysosomal export is impaired. Conversely, Spd supplementation, administered at early or later stages, reverses neuroinflammation, and improves motor and non-motor phenotypes.

In contrast to Spd, Spm injection exacerbates toxicity in *Atp13a2* knockout mice, and oral Spm supplementation fails to restore behavioral outcomes despite increasing systemic and brain polyamine levels. Spm supplementation increases microglial lysosomal abnormalities, in line with previous reports linking elevated Spm to impaired lysosomal integrity and ion channel excitability^10,72,73^. On the contrary, Spd supplementation restores microglia lysosomal deficits, pointing to a differential impact of polyamine species on lysosomal health. Our findings illustrate that total brain polyamine levels are insufficient to explain disease phenotypes, but instead highlight the importance of subcellular polyamine distribution downstream the lysosome as well as polyamine species effects.

Together, ATP13A2 represents a central regulator of brain polyamine homeostasis. ATP13A2 deficiency leads to brain polyamine shortage that emerges as an early contributor rather than a downstream consequence of neuroinflammation and parkinsonian behavioural symptoms, which can be reversed by Spd supplementation. This positions ATP13A2 at the center of a lysosome-mitochondria-inflammation axis via lysosomal polyamine transport.

### ATP13A3 enables spermidine-dependent rescue of glial inflammatory state

We identify Spd as a key modulator of the inflammatory response downstream of ATP13A2 loss. Spd supplementation rescues lysosomal and mitochondrial dysfunction in microglia, suppresses NF-κB activation, and reduces the release of neurotoxic cytokines, thereby restoring neuronal integrity in co-culture systems. These findings are consistent with previous reports showing that Spd exerts anti-oxidant effects as well as potent anti-inflammatory properties in macrophages and microglia through inhibition of NF-κB signaling and suppression of inflammasome activation^74–76^. Spd emerges as a critical determinant of glial immune state within the brain, which may be relevant for several forms of neurodegeneration.

A key mechanistic insight from this study is the identification of ATP13A3 as a compensatory polyamine transporter that enables Spd-mediated rescue in the absence of ATP13A2. We show that inhibition or silencing of ATP13A3 abolishes the anti-inflammatory effects of Spd in microglia, demonstrating that cellular uptake is required for its protective function. The upregulation of ATP13A3 in ATP13A2-deficient microglia suggests an adaptive response aimed at restoring polyamine homeostasis.

Our study provides a mechanistic framework for how exogenous Spd can bypass defective lysosomal export and exert beneficial intracellular effects, and positions ATP13A3 as a potential therapeutic entry point for modulating glial function.

### Therapeutic implications, limitations and future directions

Collectively, our findings identify early brain polyamine deficiency as a modifiable contributor to ATP13A2-associated parkinsonism and demonstrate that Spd supplementation can modulate this pathway across multiple models reversing neuroinflammation in mice as well as neurodegeneration in flies. Our results provide a strong rationale for exploring polyamine-based interventions in Kufor-Rakeb syndrome, and suggests gliosis is central to ATP13A2-deficiency, although evidence in human patients is restricted to one case report. Spd-mediated rescue of neurodegeneration in multiple PD fly models further suggests broader applications in defined subsets of PD. Given that lysosomal dysfunction and neuroinflammation are shared features across genetic and sporadic PD, our observations raise the possibility that disrupted polyamine homeostasis represents a broader and previously unrecognized contributor to PD pathogenesis. Altered polyamine profiles have been reported in the plasma and cerebrospinal fluid of patients with sporadic PD, and reduced ATP13A2 expression has been observed in dopaminergic neurons^9,18,19^. Several polyamine-modulating therapeutic interventions may be used to treat Kufor-Rakeb syndrome, such as activation of ATP13A3 and/or Spd supplementation, whereas in other parkinsonian disorders, ATP13A2 agonists may also be considered.

This work supports further exploration of Spd supplementation as a therapeutic strategy for ATP13A2 deficiency. Spd supplements are on the market, and doses as high as 40 mg/day are documented to be safe in healthy individuals^77^. However, careful evaluation of dosing, brain penetration, and long-term effects will be required. Future studies need to further define optimal dosing, treatment windows, and patient stratification, as well as to further resolve neuron-intrinsic *versus* glia-mediated effects *in vivo*. In addition, the precise molecular pathways linking polyamine depletion to inflammatory gene programs warrant further investigation, although our parallel study suggests that metabolic epigenetic reprogramming as a result of polyamine deficiency may be a key underlying factor^64^. Finally, whether similar polyamine-dependent mechanisms operate in sporadic PD or other monogenetic PD forms requires future validation.

In conclusion, we establish polyamine homeostasis as a central regulator of glial inflammatory state and neuronal vulnerability, providing a mechanistic framework that links lysosomal dysfunction to metabolic polyamine shortage and neuroinflammation, and identifies new avenues for therapeutic intervention in Kufor-Rakeb syndrome and/or PD.

## DECLARATIONS

Animal experiments were carried out in accordance with the European Communities Council Directive of November 24, 1986 (86/609/EEC) and approved by the Bioethical Committee of the KU Leuven (Belgium) (ECD project 69/2021). The patient was initially examined in Lübeck under ethics approval AZ07-113 (Genepark) (which was without post mortem analyses). In the federal state of Hamburg, Germany, using anonymized specimens from autopsies for research is permitted (Hamburg Hospital Act, Hamburgisches Krankenhausgesetz HmbKHG §12). All alive brothers and sister of the patient gave written consent to perform a brain autopsy in June 2020.

## DATA AVAILABILITY

All data, protocols and lab material are listed on the Key Resource Table with the respective DOI or identifiers at https://10.5281/zenodo.19672835. All data generated or analysed in this study can be found through the Zenodo repository at DOI: 10.5281/zenodo.19672835. All protocols are shared via protocols.io at (https://www.protocols.io/edit/cascalho-sati-et-aljwb6cparf). All data and protocols will be made publicly available as of the date of publication.

## COMPETING INTERESTS

For the sake of transparency, it seems appropriate to declare that Pe.Va. is a scientific co-founder of EndLyz Therapeutics. The company had no role in study design, data collection, data analysis, decision to publish, or preparation of the manuscript. C. K. has served as a medical advisor to Centogene and Biogen, and received speakers’ honoraria from Bial and royalties from Oxford University Press and Springer Nature

## FUNDING

This research was funded by the Aligning Science Across Parkinson’s [ASAP-024297] through the Michael J. Fox Foundation for Parkinson’s Research (MJFF) allocated to Pe.Va., V.B., J.B. and Pa.Ve. Further support comes from the FWO research grants (G094219N and G009324N), the C1 KU Leuven research grants (C14/21/095 and C14/25/154) and the C3 KU Leuven research grant (C3/22/048) assigned to Pe.Va. For the purpose of open access, the author has applied a CC BY public copyright license to all Author Accepted Manuscripts arising from this submission.

## AUTHORS’ CONTRIBUTIONS

The study was designed and coordinated by A.C., A.S. and Pe.Va.; A.C., A.S., H.D., N.S. and A.M. performed and analysed mouse behaviour experiments and polyamine modulation treatments; A.C. and A.S. applied for ethical approval; A.S., H.D. and N.S. analysed neuroinflammation; A.S. and H.D. isolated and cultured primary microglia (with/without neurons); N.K. and Pa.Ve. designed and performed Drosophila experiments; E.C. and J.B. performed and analysed human iPSC-derived microglia studies; M. B., M.G., N.B. and C.K provided and performed neuropathological examination of human patient sample. V.B. provided infrastructure for primary cell isolations; J.E. and J.B. offered valuable discussions; Pe.Va. provided main funding for the study; A.S. and Pe.Va. wrote the manuscript, which was edited and approved by all authors.

## Supporting information

Supplementary Table 1_key resources table

## ACKNOWLEDGEMENTS

We acknowledge the VIB Bioimaging Core for the use of the confocal microscope and technical assistance. Metabolomics Expertise Center (VIB-KU Leuven) for performing all the metabolomics analysis. Cell and Tissue Imaging Cluster (supported by Hercules AKUL/15/37_GOH1816N and FWO G.0929.15 to P. Vanden Berghe, KU Leuven).

## DECLARATION OF GENERATIVE AI AND AI-ASSISTED TECHNOLOGIES IN THE MANUSCRIPT PREPARATION PROCESS

During the preparation of this work the authors used ChatGPT (OpenAI) in order to screen for errors in the manuscript and assist with editing. After using this tool/service, the authors reviewed and edited the content as needed and take full responsibility for the content of the published article.

**Supplementary Figure 1. Atp13a2 loss drives early parkinsonian deficits and changes in polyamine levels.** (a) Immobility time of *Atp13a2* WT and KO mice. 4 weeks, N = 13 WT and 11 KO mice; 10 weeks, N = 19 WT and 17 KO mice; 6 months, N = 17 WT and 17 KO mice. 12 months, N = 13 WT and 11 KO mice. (b) Freezing time of *Atp13a2* WT and KO. 4 weeks, N = 12 WT and 11 KO mice; 10 weeks, N = 20 WT and 17 KO mice; 6 months, N = 17 WT and 17 KO mice. 12 months, N = 13 WT and 11 KO mice. (c) Number of faecal pellets produced by *Atp13a2* WT and KO mice at 10 weeks of age. N = 18 WT and 15 KO mice. (d) Percentage moisture of faecal matter produced at 10 weeks of age. N = 18 WT and 15 KO mice. (e) Representative images of cortical microglia (Iba-1, black) in PND14 *Atp13a2* WT and KO brains. Scale bar = 200 µm. (f) Iba-1 positive area in the cortex of PND14 brains. N = 3 WT and 3 KO mice. (g) The area positive for Iba-1 in the striatum. N = 3 WT and 3 KO mice. (h) The area positive for Iba-1 in the cerebellum. N = 3 WT and 3 KO mice. (i) GFAP positive area in the cortex. N = 3 WT and 3 KO mice. (j) The area positive for GFAP in the striatum. N = 3 WT and 3 KO mice. (k) The area positive for GFAP in the cerebellum. N = 3 WT and 3 KO mice. (l) Iba-1 positive area in the striatum at 10 weeks of age. N = 4 WT and 5 KO mice. (m) The area positive for Iba-1 in the cerebellum at 10 weeks of age. N = 4 WT and 5 KO mice. (n) Representative images of cortical astrocytes (GFAP, black) in 10-week-old *Atp13a2* WT and KO brains. Scale bar = 200 µm. (o) GFAP positive area in the cortex. (p) The area positive for GFAP in the striatum. N = 4 WT and 5 KO mice. (q) The area positive for GFAP in the cerebellum. N = 4 WT and 5 KO mice. (r) CD68 positive area in the cerebellum. N = 4 WT and 5 KO mice. (s) Representative of NF-κB (magenta) in the striatum of *Atp13a2* WT and KO mice. Scale bar = 20 µm. DAPI (cyan). (t) The intensity of the nuclear fraction of NF-κB immunostaining in the striatum and the cortex of *Atp13a2* WT and KO mice. N = 5 per genotype. (u) The abundance of polyamine species in the plasma of *Atp13a2* WT and KO mice at PND14. N = 13 WT and 8 KO mice. (v) The abundance of polyamine species in the plasma of 10 weeks mice. N = 9 WT and 6 KO mice. (w) The abundance of polyamine species in the liver of PND14 *Atp13a2* WT and KO mice. N = 3 per genotype. (x) Enrichment of ATP13A2 RNA expression in the human across the brain and various tissues. (y) Enrichment of ATP13A2 RNA expression across various tissues in the mouse. Mixed effects with Šídák’s multiple comparisons post-test (a-b, u-w). Unpaired Student’s t-test (c-d,m,p,r,t). Mann-Whitney test (f-k, m,o,q,j). Ornithine, Orn; Putrescine, Put; Spermidine, Spd; Spermine, Spm; N^1^-Acetylated Spermine, N^1^-AcSpm; N^1^/N^8^-Acetylated Spermidine, N^1^/N^8^-AcSpd Diagram was created on Biorender.com.

**Supplementary Figure 2. Juvenile negative polyamine modulation and polyamine supplementation with Spd and Spm have no impact on body weights, water and food intake.** (a) Body weights of *Atp13a2* WT and KO mice exposed to negative (DFMO and DenSpm) and positive (spermidine, Spd, and spermine, Spm) modulators. N = 33 WT, 58 WT (DFMO), 8 WT (DenSpm), 39 KO mice, 46 KO (DFMO) and 31 KO (DenSpm). (b) Average water intake of one mouse per cage of DFMO treated *Atp13a2* WT and KO mice. N = 8 WT cages, 12 KO cages, 11-15 WT (DFMO) and 11 KO (DFMO). (c) Immobility time of *Atp13a2* WT and KO mice treated with DFMO or DenSpm in the open field at 10 weeks of age. N = 18 WT, 25 WT (DFMO), 13 WT (DenSpm), 28 KO mice, 7 KO (DFMO) and 20 KO (DenSpm). (d) Freezing time of *Atp13a2* WT and KO mice treated with DFMO or DenSpm in the open field at 10 weeks of age. N = 24 WT, 25 WT (DFMO), 13 WT (DenSpm), 28 KO mice, 7 KO (DFMO) and 20 KO (DenSpm). (e) Body weights of *Atp13a2* WT and KO mice supplemented with Spd and Spm from PND2 to 10 weeks of age. N = 16 WT, 20 WT (Spm), 9 WT (Spd), 11 KO mice, 12 KO (Spm) and 9 KO (Spd). (f) Average water intake calculated for one mouse per cage of *Atp13a2* WT and KO mice given Spd or Spm supplementation from weaning until 10 weeks of age. N = 13 WT cages, 11 KO cages, 11 WT (Spm), 5 WT (Spd), KO mice, 10 KO (Spm) and 6 KO (Spd). (g) Average food intake calculated for one mouse per cage of *Atp13a2* WT and KO mice with Spd or Spm supplementation from PND2 to 10 weeks of age. N = 13 WT cages, 11 KO cages, 11 WT (Spm), 5 WT (Spd), KO mice, 10 KO (Spm) and 6 KO (Spd). (h) Freezing time of *Atp13a2* WT and KO mice treated with Spd or control in the open field at 12 months. N = 13 WT, 15 KO, 5 WT (Spd) and 5 KO (Spd). (i) Speed of *Atp13a2* WT and KO mice treated with Spd or control at 12 months. N = 16 WT, 15 KO, 5 WT (Spd) and 6 KO (Spd). Data are represented as mean ± SEM. RM Two-way ANOVA (a,e). Mixed effects with Tukey’s multiple comparisons post-test (b,f,g). One-way ANOVA with Bonferroni multiple comparisons test (c-d). One-way ANOVA with Bonferroni (h) and Holm-Šídák’s (i) multiple comparisons post-test.

**Supplementary Figure 3. Juvenile spermine supplementation does not rescue motor and non-motor defects or neuroinflammation in *Atp13a2* KO mice.** (a) Timeline for spermine (Spm) supplementation in *Atp13a2* WT and KO mice. (b) The percentage survival of *Atp13a2* WT and KO mice treated with Spm or control. N = 16 WT, 12 KO, 24 WT (Spm) and 12 KO (Spm). (c) Heatmaps depicting preferred zones by *Atp13a2* WT and KO mice with Spm supplementation in the open field at 12 months. (d) The total distance traversed by Spm treated *Atp13a2* WT and KO mice in the open field at 10 weeks, 6 and 12 months. 10 weeks, N = 19 WT, 17 KO, 15 WT (Spm) and 15 KO (Spm). 6 months, N = 19 WT, 17 KO, 16 WT (Spm) and 16 KO (Spm). 12 months, N = 13 WT, 12 KO, 14 WT (Spm) and 10 KO (Spm). (e) Immobility time of *Atp13a2* WT and KO mice treated with Spm in the open field at 10 weeks, 6 and 12 months. 10 weeks, N = 19 WT, 17 KO, 15 WT (Spm) and 17 KO (Spm). 6 months, N = 17 WT, 17 KO, 16 WT (Spm) and 16 KO (Spm). 12 months, N = 13 WT, 11 KO, 14 WT (Spm) and 10 KO (Spm). (f) Freezing time of *Atp13a2* WT and KO mice supplemented with Spm in the open field at 12 months. N = 13 WT, 15 KO, 15 WT (Spm) and 10 KO (Spm). (g) Speed of *Atp13a2* WT and KO mice treated with Spm in the open field at 12 months. N = 16 WT, 15 KO, 14 WT (Spm) and 15 KO (Spm). (h) Time spent by Spm treated *Atp13a2* WT and KO in the centre and periphery of the open field at 10 weeks, 6 and 12 months. 10 weeks, N = 10 WT, 17 KO, 16 WT (Spm) and 17 KO (Spm). 6 months, N = 17 WT, 17 KO, 16 WT (Spm) and 16 KO (Spm). 12 months, N = 13 WT, 11 KO, 14 WT (Spm) and 9 KO (Spm). (i) Schematic illustrating the buried pellet test. (j) Time taken to locate the pellet in the buried pellet test by female Spm treated *Atp13a2* WT and KO mice at 10 weeks of age. N = 9 WT, 13 KO, 9 WT (Spm) and 7 KO (Spm). (k) Time taken to locate the pellet in the buried pellet test by male Spm treated *Atp13a2* WT and KO mice at 10 weeks of age. N = 10 WT, 6 KO, 6 WT (Spm) and 9 KO (Spm). (l) The abundance of polyamine species in the brain of *Atp13a2* WT and KO mice treated with Spm at PND14. N = 5 WT, 4 KO, 4 WT (Spm) and 4 KO (Spm). (m) The abundance of polyamine species in the plasma of Spm treated *Atp13a2* WT and KO at PND14. N = 7 WT, 4 KO, 3 WT (Spm) and 4 KO (Spm). Data are represented as mean ± SEM. Log-rank test (b). Two-way ANOVA with Bonferroni multiple comparisons post-test (d,h). One-way ANOVA with Tukey’s post-test (f,g,j,k). Mixed effects analysis with Tukey’s post-test (l-m). Ornithine, Orn; Putrescine, Put; Spermidine, Spd; Spermine, Spm; N^1^-Acetylated Spermine, N^1^-AcSpm; N^1^/N^8^-Acetylated Spermidine, N^1^/N^8^-AcSpd. Illustration was created on Biorender.com.

**Supplementary Figure 4. Juvenile spermine supplementation does not reverse signs of neuroinflammation in the *Atp13a2* KO mice.** (a) Representative images of cortical astrocytes (GFAP, black) of *Atp13a2* WT and KO mice treated with Spermidine (Spd). Scale bar = 200 µm. (b) The area positive for GFAP in the cortex *Atp13a2* WT and KO mice treated with Spd. N = 4 WT and 5 KO, 7 WT (Spd) and 4 KO (Spd). (c) GFAP positive area in the cerebellum. N = 4 WT and 5 KO, 7 WT (Spd) and 4 KO (Spd). (d) The area positive for GFAP striatum. N = 4 WT and 5 KO, 7 WT (Spd) and 4 KO (Spd). (e) Representative images of cortical astrocytes (GFAP, black) within the cortex of *Atp13a2* WT and KO mice supplemented with spermine (Spm). Scale bar = 200 µm. (f) The area positive for GFAP in the cortex of mice supplemented with Spm. N = 4 WT and 5 KO, 6 WT (Spm), 5 KO (Spm). (g) The area positive for GFAP in the cerebellum. (h) The area positive for GFAP in the striatum. N = 4 WT and 5 KO, 6 WT (Spm), 5 KO (Spm). (i) Representative images of cortical microglia (Iba-1, black) of *Atp13a2* WT and KO mice treated with Spm compared to vehicle control. Scale bar = 200 µm. (j) The area positive for Iba-1 within the cortex of 10-weeks-old *Atp13a2* WT and KO mice given Spm. N = 4 WT and 5 KO, 6 WT (Spm), 5 KO (Spm). (k) The area positive for Iba-1 in the cerebellum. N = 4 WT and 5 KO, 6 WT (Spm), 5 KO (Spm). (l) The area positive for Iba-1 in the striatum. N = 4 WT and 5 KO, 6 WT (Spm), 5 KO (Spm). (m) Representative images of cortical microglia (CD68, white) of *Atp13a2* WT and KO mice with Spm supplementation. Scale bar = 200 µm. (n) The area positive for CD68 in the cortex of 10-weeks-old *Atp13a2* WT and KO mice with Spm supplementation. N = 4 WT and 5 KO, 6 WT (Spm), 5 KO (Spm). (o) The area positive for CD68 in cerebellum. N = 4 WT and 5 KO, 6 WT (Spm), 5 KO (Spm). (p) Representative of NF-κB (magenta) in the striatum of *Atp13a2* WT and KO mice with Spd supplementation. Scale bar = 20 µm. DAPI (cyan). (q) Intensity of nuclear NF-κB in the striatum. N = 5 WT and 5 KO, 5 WT (Spd) and 4 KO (Spd). (r) Intensity of nuclear NF-κB in the cortex. N = 5 WT and 5 KO, 5 WT (Spd) and 4 KO (Spd). (s) Representative images of Lamp1 (yellow) and Galectin-3 (Gal-3, white) within striatal microglia (Iba-1, magenta) of *Atp13a2* WT and KO mice treated with Spm or vehicle control. Scale bar = 10 µm. (t) Lamp1 volume per microglia volume of *Atp13a2* WT and KO striatum. N = 4 WT and 5 KO, 5 WT (Spd) and 3 KO (Spd). (u) Volume of Gal-3 co-localised with Lamp1 per microglia volume. N = 4 WT and 5 KO, 5 WT (Spd) and 3 KO (Spd). Data are represented as mean ± SEM. One-way ANOVA with Bonferroni (b), Šídák’s (c-d,f-h,j-l,n-o,u) and Tukey’s (q-r,t) multiple comparisons post-tests.

**Supplementary Figure 5. Spermidine mediates anti-inflammatory and anti-oxidant effects in microglia and improves neuronal morphology.** (a) Illustration of lipopolysaccharide (LPS) and Spermidine (Spd) treatments of isolated *Atp13a2* WT and KO microglia. (b-d) The concentrations of IL-6 (b), CXCL1 (c) and IL-1β (d) in the supernatant of *Atp13a2* WT and KO microglia supplemented with LPS and Spd. N = 3 WT, 3 KO, 3 WT (LPS), 3 KO (LPS), 3 WT (0.3 µM + LPS), 3 KO (0.3 µM + LPS), 2 WT (0.3 µM Spd) and 2 KO (0.3 µM Spd). (e) Detecting mitochondrial reactive oxidative species (ROS) in *Atp13a2* WT and KO microglia supplemented with combinations of LPS, mitoTEMPO (MT) and Spd. (f) Maximal projections of MitoSox (magenta) in *Atp13a2* WT and KO microglia. DAPI (cyan); Scale bar = 50 µm. (g) Mean intensity of MitoSox in *Atp13a2* WT and KO microglia treated with a combination of LPS, Spd and MT. N = 11 WT, 11 KO, 12 WT (LPS), 2 WT (0.3 µM Spd), 2 KO (0.3 µM Spd), 2 KO (0.05 µM MT), 2 WT (1 µM MT), 2 KO (1 µM MT), 2 WT (0.05 µM MT + 0.3 µM Spd), 2 KO (0.05 µM MT + 0.3 µM Spd), 2 WT (1 µM MT + 0.3 µM Spd), 2 KO (1 µM MT), 10 KO (LPS), 11 WT (0.3 µM Spd + LPS), 12 KO (0.3 µM Spd + LPS), 9 WT/KO (LPS + 0.05 µM MT), 9 WT/KO (LPS + 1 µM MT), 9 WT/KO (LPS + 0.05 µM MT + Spd), 9 WT/KO (LPS + 1 µM MT + Spd). (h) Spd treatment in microglia and subsequent establishment of microglia and neurons co-cultures. (i) Representative images of *Atp13a2* WT neurons co-cultured with Spd-treated and untreated *Atp13a2* KO microglia. DAPI (cyan); Scale bar = 20 µm. (j) Neurite length of *Atp13a2* WT neurons in co-cultures with Spd-treated and untreated *Atp13a2* KO and WT microglia. N = 3 per treatment. Data are represented as mean ± SEM. Mixed effects analysis with Tukey’s post-test (b-d,g,j).

## Notes

### Author Declarations

Ethics Committee (EC) of the University of Lubeck, Germany gave the ethics approval (number AZ07-113; Genepark) for the patient assessment (which was without postmortem). The federal state of Hamburg, Germany, approved the use of the autopsy research under the the Hamburg Hospital Act, Hamburgisches Krankenhausgesetz HmbKHG 12.

## References

1. Sikora, J., Dovero, S., Kinet, R., Arotcarena, M.L., Bohic, S., Bezard, E., Fernagut, P.O., and Dehay, B. (2024). Nigral ATP13A2 depletion induces Parkinson’s disease-related neurodegeneration in a pilot study in non-human primates. NPJ Parkinsons Dis 10, 141. 10.1038/s41531-024-00757-4.

2. Kett, L.R., Stiller, B., Bernath, M.M., Tasset, I., Blesa, J., Jackson-Lewis, V., Chan, R.B., Zhou, B., Di Paolo, G., Przedborski, S., et al. (2015). α-Synuclein-independent histopathological and motor deficits in mice lacking the endolysosomal Parkinsonism protein Atp13a2. J Neurosci 35, 5724–5742. 10.1523/jneurosci.0632-14.2015.

3. Ramirez, A., Heimbach, A., Gründemann, J., Stiller, B., Hampshire, D., Cid, L.P., Goebel, I., Mubaidin, A.F., Wriekat, A.L., Roeper, J., et al. (2006). Hereditary parkinsonism with dementia is caused by mutations in ATP13A2, encoding a lysosomal type 5 P-type ATPase. Nat Genet 38, 1184–1191. 10.1038/ng1884.

4. Di Fonzo, A., Chien, H.F., Socal, M., Giraudo, S., Tassorelli, C., Iliceto, G., Fabbrini, G., Marconi, R., Fincati, E., Abbruzzese, G., et al. (2007). ATP13A2 missense mutations in juvenile parkinsonism and young onset Parkinson disease. Neurology 68, 1557–1562. 10.1212/01.wnl.0000260963.08711.08.

5. Estrada-Cuzcano, A., Martin, S., Chamova, T., Synofzik, M., Timmann, D., Holemans, T., Andreeva, A., Reichbauer, J., De Rycke, R., Chang, D.I., et al. (2017). Loss-of-function mutations in the ATP13A2/PARK9 gene cause complicated hereditary spastic paraplegia (SPG78). Brain 140, 287–305. 10.1093/brain/aww307.

6. Bras, J., Verloes, A., Schneider, S.A., Mole, S.E., and Guerreiro, R.J. (2012). Mutation of the parkinsonism gene ATP13A2 causes neuronal ceroid-lipofuscinosis. Hum Mol Genet 21, 2646–2650. 10.1093/hmg/dds089.

7. Yang, X., and Xu, Y. (2014). Mutations in the ATP13A2 gene and Parkinsonism: a preliminary review. Biomed Res Int 2014, 371256. 10.1155/2014/371256.

8. Chien, H.F., Rodriguez, R.D., Bonifati, V., Nitrini, R., Pasqualucci, C.A., Gelpi, E., and Barbosa, E.R. (2021). Neuropathologic Findings in a Patient With Juvenile-Onset Levodopa-Responsive Parkinsonism Due to ATP13A2 Mutation. Neurology 97, 763–766. doi:10.1212/WNL.0000000000012705.

9. Dehay, B., Ramirez, A., Martinez-Vicente, M., Perier, C., Canron, M.H., Doudnikoff, E., Vital, A., Vila, M., Klein, C., and Bezard, E. (2012). Loss of P-type ATPase ATP13A2/PARK9 function induces general lysosomal deficiency and leads to Parkinson disease neurodegeneration. Proc Natl Acad Sci U S A 109, 9611–9616. 10.1073/pnas.1112368109.

10. van Veen, S., Martin, S., Van den Haute, C., Benoy, V., Lyons, J., Vanhoutte, R., Kahler, J.P., Decuypere, J.-P., Gelders, G., Lambie, E., et al. (2020). ATP13A2 deficiency disrupts lysosomal polyamine export. Nature 578, 419–424. 10.1038/s41586-020-1968-7.

11. Vrijsen, S., Besora-Casals, L., van Veen, S., Zielich, J., Van den Haute, C., Hamouda, N.N., Fischer, C., Ghesquière, B., Tournev, I., Agostinis, P., et al. (2020). ATP13A2-mediated endo-lysosomal polyamine export counters mitochondrial oxidative stress. Proc Natl Acad Sci U S A 117, 31198–31207. 10.1073/pnas.1922342117.

12. Igarashi, K., and Kashiwagi, K. (2010). Modulation of cellular function by polyamines. Int J Biochem Cell Biol 42, 39–51. 10.1016/j.biocel.2009.07.009.

13. Vrijsen, S., Houdou, M., Cascalho, A., Eggermont, J., and Vangheluwe, P. (2023). Polyamines in Parkinson’s Disease: Balancing Between Neurotoxicity and Neuroprotection. Annual Review of Biochemistry 92, 435–464. 10.1146/annurev-biochem-071322-021330.

14. Usenovic, M., Tresse, E., Mazzulli, J.R., Taylor, J.P., and Krainc, D. (2012). Deficiency of ATP13A2 leads to lysosomal dysfunction, α-synuclein accumulation, and neurotoxicity. J Neurosci 32, 4240–4246. 10.1523/jneurosci.5575-11.2012.

15. Tsunemi, T., Perez-Rosello, T., Ishiguro, Y., Yoroisaka, A., Jeon, S., Hamada, K., Rammonhan, M., Wong, Y.C., Xie, Z., Akamatsu, W., et al. (2019). Increased Lysosomal Exocytosis Induced by Lysosomal Ca(2+) Channel Agonists Protects Human Dopaminergic Neurons from α-Synuclein Toxicity. J Neurosci 39, 5760–5772. 10.1523/jneurosci.3085-18.2019.

16. Grünewald, A., Arns, B., Seibler, P., Rakovic, A., Münchau, A., Ramirez, A., Sue, C.M., and Klein, C. (2012). ATP13A2 mutations impair mitochondrial function in fibroblasts from patients with Kufor-Rakeb syndrome. Neurobiol Aging 33, 1843.e1841–1847. 10.1016/j.neurobiolaging.2011.12.035.

17. Samaddar, M., Fitzgerald, G.A., Nguyen, A.H., Davis, S.S., Jain, S., Guo, J., Propson, N.E., van Lengerich, B., Shi, Y., Balasundar, S., et al. (2025). Lysosomal polyamine storage upon ATP13A2 loss impairs β-glucocerebrosidase via altered lysosomal pH and electrostatic hydrolase-lipid interactions. Cell Reports 44. 10.1016/j.celrep.2025.116179.

18. Saiki, S., Sasazawa, Y., Fujimaki, M., Kamagata, K., Kaga, N., Taka, H., Li, Y., Souma, S., Hatano, T., Imamichi, Y., et al. (2019). A metabolic profile of polyamines in parkinson disease: A promising biomarker. Annals of Neurology 86, 251–263. 10.1002/ana.25516.

19. Paik, M.-J., Ahn, Y.-H., Lee, P.H., Kang, H., Park, C.B., Choi, S., and Lee, G. (2010). Polyamine patterns in the cerebrospinal fluid of patients with Parkinson’s disease and multiple system atrophy. Clinica Chimica Acta 411, 1532–1535. 10.1016/j.cca.2010.05.034.

20. Kinet, R., Sikora, J., Arotcarena, M.L., Decourt, M., Balado, E., Doudnikoff, E., Bohic, S., Vesnaver, M., Lovisotto, A., Thiolat, M.L., et al. (2025). Phenotypic characterization of an Atp13a2 knockout rat model of Parkinson’s disease. NPJ Parkinsons Dis 11, 321. 10.1038/s41531-025-01171-0.

21. Schmutz, I., Jagannathan, V., Bartenschlager, F., Stein, V.M., Gruber, A.D., Leeb, T., and Katz, M.L. (2019). ATP13A2 missense variant in Australian Cattle Dogs with late onset neuronal ceroid lipofuscinosis. Molecular Genetics and Metabolism 127, 95–106. 10.1016/j.ymgme.2018.11.015.

22. Tyynelä, J., Cooper, J.D., Khan, M.N., Shemilt, S.J., and Haltia, M. (2004). Hippocampal Pathology in the Human Neuronal Ceroid-Lipofuscinoses: Distinct Patterns of Storage Deposition, Neurodegeneration and Glial Activation. Brain Pathology 14, 349–357. 10.1111/j.1750-3639.2004.tb00077.x.

23. Ho, M.S. (2019). Microglia in Parkinson’s Disease. Adv Exp Med Biol 1175, 335–353. 10.1007/978-981-13-9913-8_13.

24. Siew, J.J., Chen, H.M., Chen, H.Y., Chen, H.L., Chen, C.M., Soong, B.W., Wu, Y.R., Chang, C.P., Chan, Y.C., Lin, C.H., et al. (2019). Galectin-3 is required for the microglia-mediated brain inflammation in a model of Huntington’s disease. Nat Commun 10, 3473. 10.1038/s41467-019-11441-0.

25. Siew, J.J., Chen, H.-M., Chiu, F.-L., Lee, C.-W., Chang, Y.-M., Chen, H.-L., Nguyen, T.N.A., Liao, H.-T., Liu, M., Hagar, H.-T., et al. (2023). Galectin-3 aggravates microglial activation and tau transmission in tauopathy. The Journal of Clinical Investigation 134. 10.1172/JCI165523.

26. Gan, Q., Fu, X., Zhou, T., Fan, N., Nan, N., Wang, Y., Yang, Y., Gou, S., Hu, L., and Zhou, S. (2026). Mitochondrial DNA drives NLRP3-IL-1β axis activation in microglia by binding to NLRP3, leading to neurodegeneration in Parkinson’s disease models. Cell Death & Disease 17, 213. 10.1038/s41419-026-08424-7.

27. Behrens, M.I., Brüggemann, N., Chana, P., Venegas, P., Kägi, M., Parrao, T., Orellana, P., Garrido, C., Rojas, C.V., Hauke, J., et al. (2010). Clinical spectrum of Kufor-Rakeb syndrome in the Chilean kindred with ATP13A2 mutations. Mov Disord 25, 1929–1937. 10.1002/mds.22996.

28. Montine, T.J., Phelps, C.H., Beach, T.G., Bigio, E.H., Cairns, N.J., Dickson, D.W., Duyckaerts, C., Frosch, M.P., Masliah, E., Mirra, S.S., et al. (2012). National Institute on Aging-Alzheimer’s Association guidelines for the neuropathologic assessment of Alzheimer’s disease: a practical approach. Acta Neuropathol 123, 1–11. 10.1007/s00401-011-0910-3.

29. Attems, J., Toledo, J.B., Walker, L., Gelpi, E., Gentleman, S., Halliday, G., Hortobagyi, T., Jellinger, K., Kovacs, G.G., Lee, E.B., et al. (2021). Neuropathological consensus criteria for the evaluation of Lewy pathology in post-mortem brains: a multi-centre study. Acta Neuropathologica 141, 159–172. 10.1007/s00401-020-02255-2.

30. Freitag, K., Sterczyk, N., Wendlinger, S., Obermayer, B., Schulz, J., Farztdinov, V., Mülleder, M., Ralser, M., Houtman, J., Fleck, L., et al. (2022). Spermidine reduces neuroinflammation and soluble amyloid beta in an Alzheimer’s disease mouse model. J Neuroinflammation 19, 172. 10.1186/s12974-022-02534-7.

31. Maglione, M., Kochlamazashvili, G., Eisenberg, T., Rácz, B., Michael, E., Toppe, D., Stumpf, A., Wirth, A., Zeug, A., Müller, F.E., et al. (2019). Spermidine protects from age-related synaptic alterations at hippocampal mossy fiber-CA3 synapses. Scientific Reports 9, 19616. 10.1038/s41598-019-56133-3.

32. Eisenberg, T., Abdellatif, M., Schroeder, S., Primessnig, U., Stekovic, S., Pendl, T., Harger, A., Schipke, J., Zimmermann, A., Schmidt, A., et al. (2016). Cardioprotection and lifespan extension by the natural polyamine spermidine. Nature Medicine 22, 1428–1438. 10.1038/nm.4222.

33. Lim, N.K.-H., Moestrup, V., Zhang, X., Wang, W.-A., Møller, A., and Huang, F.-D. (2018). An Improved Method for Collection of Cerebrospinal Fluid from Anesthetized Mice (1940-087X). doi:10.3791/56774.

34. Tatem, K.S., Quinn, J.L., Phadke, A., Yu, Q., Gordish-Dressman, H., and Nagaraju, K. (2014). Behavioral and locomotor measurements using an open field activity monitoring system for skeletal muscle diseases. J Vis Exp, 51785. 10.3791/51785.

35. Fleming, S.M., Salcedo, J., Fernagut, P.-O., Rockenstein, E., Masliah, E., Levine, M.S., and Chesselet, M.-F. (2004). Early and Progressive Sensorimotor Anomalies in Mice Overexpressing Wild-Type Human α-Synuclein. The Journal of Neuroscience 24, 9434–9440. 10.1523/jneurosci.3080-04.2004.

36. Miedel, C.J., Patton, J.M., Miedel, A.N., Miedel, E.S., and Levenson, J.M. (2017). Assessment of Spontaneous Alternation, Novel Object Recognition and Limb Clasping in Transgenic Mouse Models of Amyloid-β and Tau Neuropathology. J Vis Exp. 10.3791/55523.

37. Lehmkuhl, A.M., Dirr, E.R., and Fleming, S.M. (2014). Olfactory assays for mouse models of neurodegenerative disease. J Vis Exp, e51804. 10.3791/51804.

38. Rota, L., Pellegrini, C., Benvenuti, L., Antonioli, L., Fornai, M., Blandizzi, C., Cattaneo, A., and Colla, E. (2019). Constipation, deficit in colon contractions and alpha-synuclein inclusions within the colon precede motor abnormalities and neurodegeneration in the central nervous system in a mouse model of alpha-synucleinopathy. Transl Neurodegener 8, 5. 10.1186/s40035-019-0146-z.

39. Bordt, E.A., Block, C.L., Petrozziello, T., Sadri-Vakili, G., Smith, C.J., Edlow, A.G., and Bilbo, S.D. (2020). Isolation of Microglia from Mouse or Human Tissue. STAR Protoc 1. 10.1016/j.xpro.2020.100035.

40. Morales-Ropero, J.M., Arroyo-Urea, S., Neubrand, V.E., Martín-Oliva, D., Marín-Teva, J.L., Cuadros, M.A., Vangheluwe, P., Navascués, J., Mata, A.M., and Sepúlveda, M.R. (2021). The endoplasmic reticulum Ca2+-ATPase SERCA2b is upregulated in activated microglia and its inhibition causes opposite effects on migration and phagocytosis. Glia 69, 842–857. 10.1002/glia.23931.

41. van Veen, S., Irala, D., Sakers, K., Savage, J., Séjourné, G., Bindu, D.S., Ausloos, E., Dhondt, H., Schoonvliet, N., Van den Haute, C., et al. (2025). Astrocytic polyamine transport by ATP13A4 tunes excitatory synaptic transmission. medRxiv, 2025.2004.2004.25325117. 10.1101/2025.04.04.25325117.

42. Azfar, M., Gao, W., Van den Haute, C., Xiao, L., Karsa, M., Pandher, R., Karsa, A., Spurling, D., Ronca, E., Bongers, A., et al. (2025). The polyamine transporter ATP13A3 mediates difluoromethylornithine-induced polyamine uptake in neuroblastoma. Mol Oncol 19, 913–936. 10.1002/1878-0261.13789.

43. Arshadi, C., Günther, U., Eddison, M., Harrington, K.I.S., and Ferreira, T.A. (2021). SNT: a unifying toolbox for quantification of neuronal anatomy. Nature Methods 18, 374–377. 10.1038/s41592-021-01105-7.

44. Stirling, D.R., Swain-Bowden, M.J., Lucas, A.M., Carpenter, A.E., Cimini, B.A., and Goodman, A. (2021). CellProfiler 4: improvements in speed, utility and usability. BMC Bioinformatics 22, 433. 10.1186/s12859-021-04344-9.

45. Kaempf, N., Valadas, J.S., Robberechts, P., Schoovaerts, N., Praschberger, R., Ortega, A., Kilic, A., Chabot, D., Pech, U., Kuenen, S., et al. (2024). Behavioral screening defines three molecular Parkinsonism subgroups in Drosophila. bioRxiv, 2024.2008.2027.609924. 10.1101/2024.08.27.609924.

46. Riemensperger, T., Issa, A.R., Pech, U., Coulom, H., Nguyễn, M.V., Cassar, M., Jacquet, M., Fiala, A., and Birman, S. (2013). A single dopamine pathway underlies progressive locomotor deficits in a Drosophila model of Parkinson disease. Cell Rep 5, 952–960. 10.1016/j.celrep.2013.10.032.

47. Maetzler, W., Liepelt, I., and Berg, D. (2009). Progression of Parkinson’s disease in the clinical phase: potential markers. The Lancet Neurology 8, 1158–1171. 10.1016/S1474-4422(09)70291-1.

48. Glass, C.K., Saijo, K., Winner, B., Marchetto, M.C., and Gage, F.H. (2010). Mechanisms Underlying Inflammation in Neurodegeneration. Cell 140, 918–934. 10.1016/j.cell.2010.02.016.

49. Nimmerjahn, A., Kirchhoff, F., and Helmchen, F. (2005). Resting microglial cells are highly dynamic surveillants of brain parenchyma in vivo. Science 308, 1314–1318. 10.1126/science.1110647.

50. Holtman, I.R., Raj, D.D., Miller, J.A., Schaafsma, W., Yin, Z., Brouwer, N., Wes, P.D., Möller, T., Orre, M., Kamphuis, W., et al. (2015). Induction of a common microglia gene expression signature by aging and neurodegenerative conditions: a co-expression meta-analysis. Acta Neuropathol Commun 3, 31 10.1186/s40478-015-0203-5.

51. Mathys, H., Adaikkan, C., Gao, F., Young, J.Z., Manet, E., Hemberg, M., De Jager, P.L., Ransohoff, R.M., Regev, A., and Tsai, L.H. (2017). Temporal Tracking of Microglia Activation in Neurodegeneration at Single-Cell Resolution. Cell Rep 21, 366–380. 10.1016/j.celrep.2017.09.039.

52. Krasemann, S., Madore, C., Cialic, R., Baufeld, C., Calcagno, N., El Fatimy, R., Beckers, L., O’Loughlin, E., Xu, Y., Fanek, Z., et al. (2017). The TREM2-APOE Pathway Drives the Transcriptional Phenotype of Dysfunctional Microglia in Neurodegenerative Diseases. Immunity 47, 566–581.e569. 10.1016/j.immuni.2017.08.008.

53. Jia, J., Claude-Taupin, A., Gu, Y., Choi, S.W., Peters, R., Bissa, B., Mudd, M.H., Allers, L., Pallikkuth, S., Lidke, K.A., et al. (2020). Galectin-3 Coordinates a Cellular System for Lysosomal Repair and Removal. Dev Cell 52, 69–87.e68. 10.1016/j.devcel.2019.10.025.

54. Yip, P.K., Carrillo-Jimenez, A., King, P., Vilalta, A., Nomura, K., Chau, C.C., Egerton, A.M., Liu, Z.H., Shetty, A.J., Tremoleda, J.L., et al. (2017). Galectin-3 released in response to traumatic brain injury acts as an alarmin orchestrating brain immune response and promoting neurodegeneration. Sci Rep 7, 41689. 10.1038/srep41689.

55. Thomas, T., and Thomas, T.J. (2001). Polyamines in cell growth and cell death: molecular mechanisms and therapeutic applications. Cell Mol Life Sci 58, 244–258. 10.1007/pl00000852.

56. Pirinen, E., Kuulasmaa, T., Pietilä, M., Heikkinen, S., Tusa, M., Itkonen, P., Boman, S., Skommer, J., Virkamäki, A., Hohtola, E., et al. (2007). Enhanced polyamine catabolism alters homeostatic control of white adipose tissue mass, energy expenditure, and glucose metabolism. Mol Cell Biol 27, 4953–4967. 10.1128/mcb.02034-06.

57. Grabenauer, M., Bernstein, S.L., Lee, J.C., Wyttenbach, T., Dupuis, N.F., Gray, H.B., Winkler, J.R., and Bowers, M.T. (2008). Spermine Binding to Parkinson’s Protein α-Synuclein and Its Disease-Related A30P and A53T Mutants. The Journal of Physical Chemistry B 112, 11147–11154. 10.1021/jp801175w.

58. Lewandowski, N.M., Ju, S., Verbitsky, M., Ross, B., Geddie, M.L., Rockenstein, E., Adame, A., Muhammad, A., Vonsattel, J.P., Ringe, D., et al. (2010). Polyamine pathway contributes to the pathogenesis of Parkinson disease. Proceedings of the National Academy of Sciences 107, 16970–16975. doi:10.1073/pnas.1011751107.

59. Gordon, R., Albornoz, E.A., Christie, D.C., Langley, M.R., Kumar, V., Mantovani, S., Robertson, A.A.B., Butler, M.S., Rowe, D.B., O’Neill, L.A., et al. (2018). Inflammasome inhibition prevents á-synuclein pathology and dopaminergic neurodegeneration in mice. Sci Transl Med 10. 10.1126/scitranslmed.aah4066.

60. Paul, B.D., Snyder, S.H., and Bohr, V.A. (2021). Signaling by cGAS-STING in Neurodegeneration, Neuroinflammation, and Aging. Trends Neurosci 44, 83–96. 10.1016/j.tins.2020.10.008.

61. Nazmi, A., Field, R.H., Griffin, E.W., Haugh, O., Hennessy, E., Cox, D., Reis, R., Tortorelli, L., Murray, C.L., Lopez-Rodriguez, A.B., et al. (2019). Chronic neurodegeneration induces type I interferon synthesis via STING, shaping microglial phenotype and accelerating disease progression. Glia 67, 1254–1276. 10.1002/glia.23592.

62. West, A.P., Khoury-Hanold, W., Staron, M., Tal, M.C., Pineda, C.M., Lang, S.M., Bestwick, M., Duguay, B.A., Raimundo, N., MacDuff, D.A., et al. (2015). Mitochondrial DNA stress primes the antiviral innate immune response. Nature 520, 553–557. 10.1038/nature14156.

63. Hamouda, N.N., Van den Haute, C., Vanhoutte, R., Sannerud, R., Azfar, M., Mayer, R., Cortés Calabuig, Á., Swinnen, J.V., Agostinis, P., Baekelandt, V., et al. (2021). ATP13A3 is a major component of the enigmatic mammalian polyamine transport system. J Biol Chem 296, 100182. 10.1074/jbc.RA120.013908.

64. Coccia, E., Parfitt, G.M., Ijaz, S., Sati, A., Gesner, J., Arevalo, A.P., Strong, J., Bright, A., Sohail, S., Meimoun, T., et al. (2026). ATP13A2 Loss of Function-Driven Polyamine Dysregulation Induces SAM Depletion and Epigenetic Astrocyte Toxicity. bioRxiv, 2026.2004.2002.716164. 10.64898/2026.04.02.716164.

65. Qiao, C., Yin, N., Gu, H.-Y., Zhu, J.-L., Ding, J.-H., Lu, M., and Hu, G. (2016). Atp13a2 Deficiency Aggravates Astrocyte-Mediated Neuroinflammation via NLRP3 Inflammasome Activation. CNS Neuroscience & Therapeutics 22, 451–460. 10.1111/cns.12514.

66. Zou, Z., Zhou, J., Lu, Y., Huang, Y., Zhou, L., Wang, X., Chen, J., Tian, H., Ge, X., Guo, C., et al. (2026). ATP13A2 restrains macrophage NLRP3 inflammasome activation to repress neurodegeneration via modulating mitochondrial homeostasis. Proceedings of the National Academy of Sciences 123, e2534066123. doi:10.1073/pnas.2534066123.

67. Sati, A., Prescott, M., Holland, S., Jasoni, C.L., Desroziers, E., and Campbell, R.E. (2021). Morphological evidence indicates a role for microglia in shaping the PCOS-like brain. Journal of Neuroendocrinology 33, e12999. 10.1111/jne.12999.

68. Marangoni, M., Adalbert, R., Janeckova, L., Patrick, J., Kohli, J., Coleman, M.P., and Conforti, L. (2014). Age-related axonal swellings precede other neuropathological hallmarks in a knock-in mouse model of Huntington’s disease. Neurobiol Aging 35, 2382–2393. 10.1016/j.neurobiolaging.2014.04.024.

69. Serrano-Pozo, A., Frosch, M.P., Masliah, E., and Hyman, B.T. (2011). Neuropathological alterations in Alzheimer disease. Cold Spring Harb Perspect Med 1, a006189. 10.1101/cshperspect.a006189.

70. Kordower, J.H., Olanow, C.W., Dodiya, H.B., Chu, Y., Beach, T.G., Adler, C.H., Halliday, G.M., and Bartus, R.T. (2013). Disease duration and the integrity of the nigrostriatal system in Parkinson’s disease. Brain 136, 2419–2431. 10.1093/brain/awt192.

71. Croucher, K., Lepp, J.K., Bechtold, J., Hamad, E.J., Scott, S., Bittner, C., Rogers, S., Ong, C., Boehme, S., Wang, Z., et al. (2025). Phenotype Differences Between ATP13A2 Heterozygous and Knockout Mice Across Aging. International Journal of Molecular Sciences 26, 7030.

72. Soulet, D., and Rivest, S. (2003). Polyamines play a critical role in the control of the innate immune response in the mouse central nervous system. J Cell Biol 162, 257–268. 10.1083/jcb.200301097.

73. Williams, K. (1997). Interactions of polyamines with ion channels. Biochem J 325 ( *Pt 2*), 289–297. 10.1042/bj3250289.

74. Yuan, H., Peng, F., Zhou, Y.F., Wu, S.X., Long, Z.Y., Peng, Y.M., and Zhou, Y.Z. (2026). Inhibitory effects of spermidine on lipopolysaccharide-induced inflammation in RAW 264.7 cells. Exp Ther Med 31, 124. 10.3892/etm.2026.13119.

75. Liu, R., Cao, L., Zhou, Y., Li, Y., Wang, T., Meng, Z., Yang, T., Feng, C., Yang, Q., Liu, X., et al. (2025). Spermidine enhances macrophages anti-inflammatory and regenerative functions by improving mitochondrial fitness. Phytomedicine 148, 157349. 10.1016/j.phymed.2025.157349.

76. Li, X., Zhou, X., Liu, X., Li, X., Jiang, X., Shi, B., and Wang, S. (2022). Spermidine protects against acute kidney injury by modulating macrophage NLRP3 inflammasome activation and mitochondrial respiration in an eIF5A hypusination-related pathway. Mol Med 28, 103. 10.1186/s10020-022-00533-1.

77. Keohane, P., Everett, J.R., Pereira, R., Cook, C.M., Blonquist, T.M., and Mah, E. (2024). Supplementation of spermidine at 40 mg/day has minimal effects on circulating polyamines: An exploratory double-blind randomized controlled trial in older men. Nutrition Research 132, 1–14. 10.1016/j.nutres.2024.09.012.

