## Supplementary figures and images for "Spermidine suppresses glial inflammation and parkinsonian abnormalities in ATP13A2 deficiency"

### Supplementary Figure 1

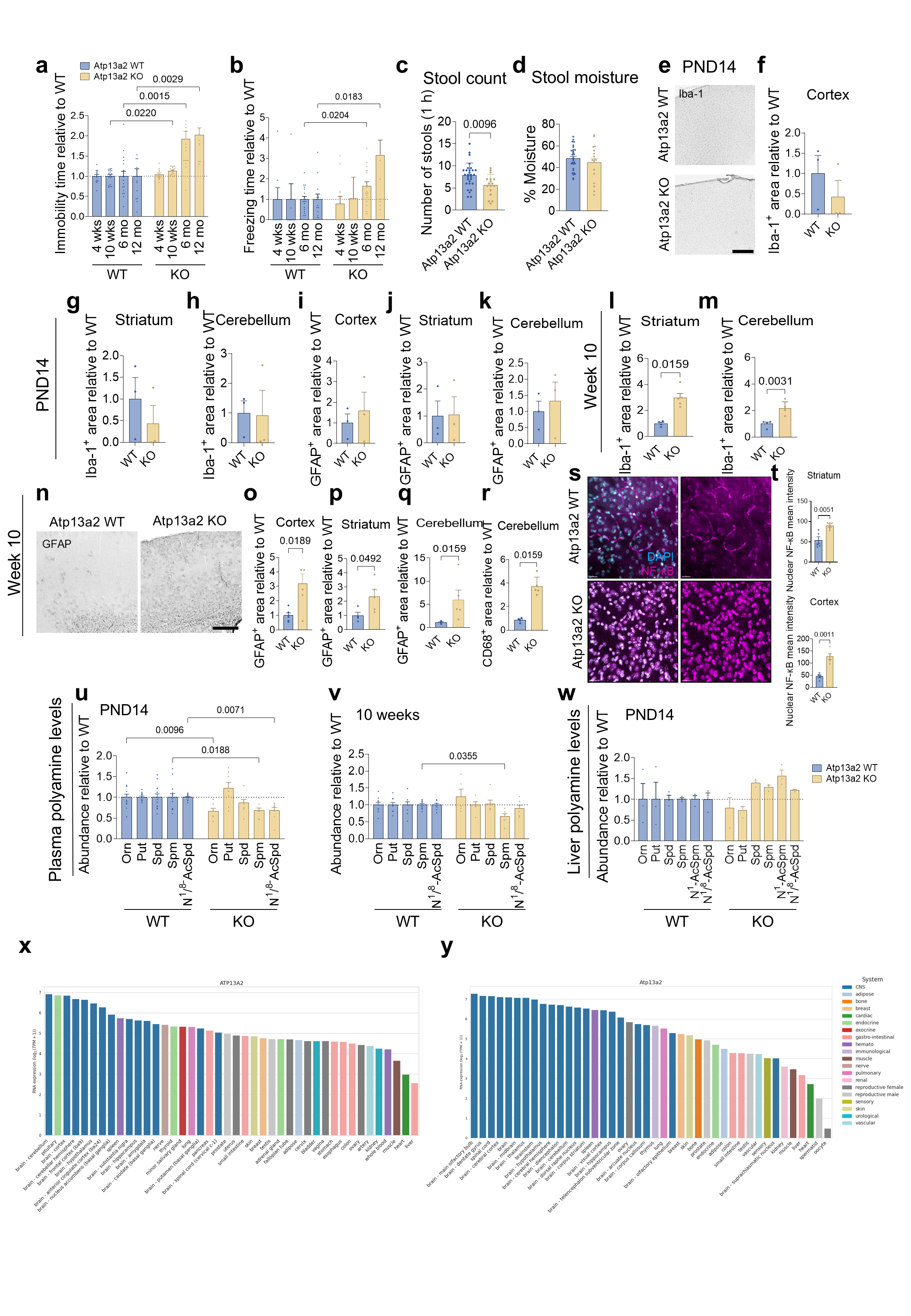

### Supplementary Figure 2

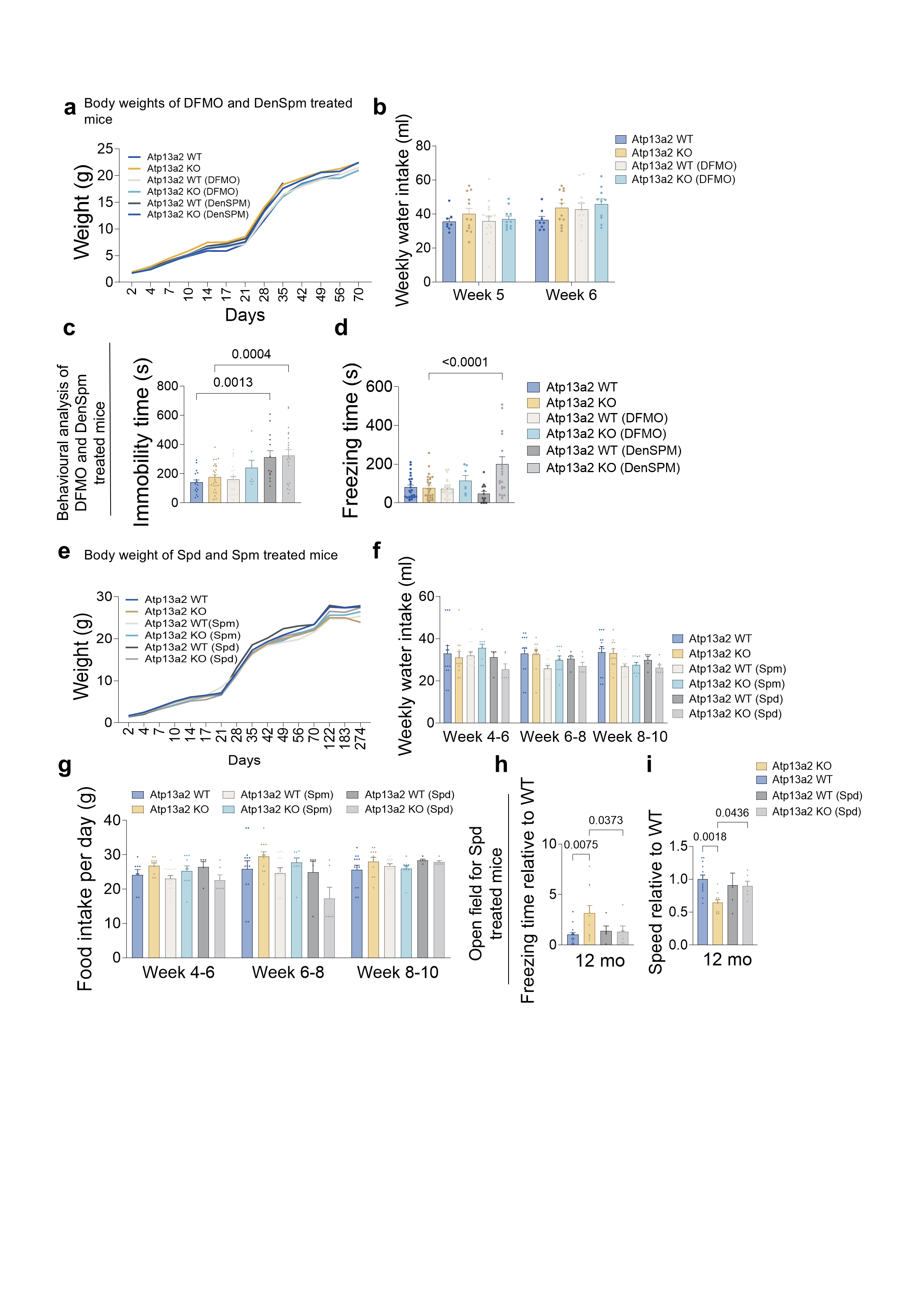

### Supplementary Figure 3

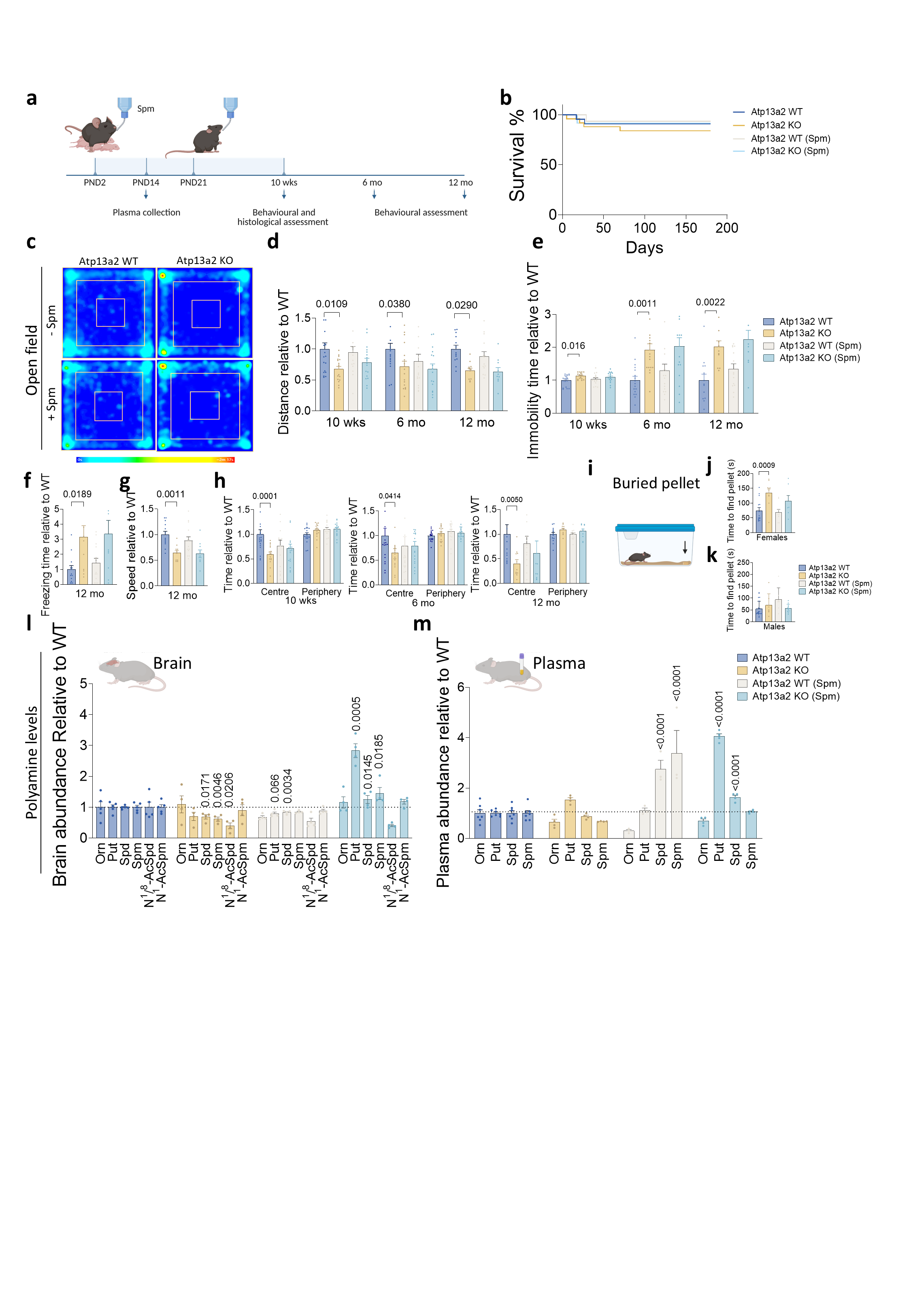

### Supplementary Figure 4

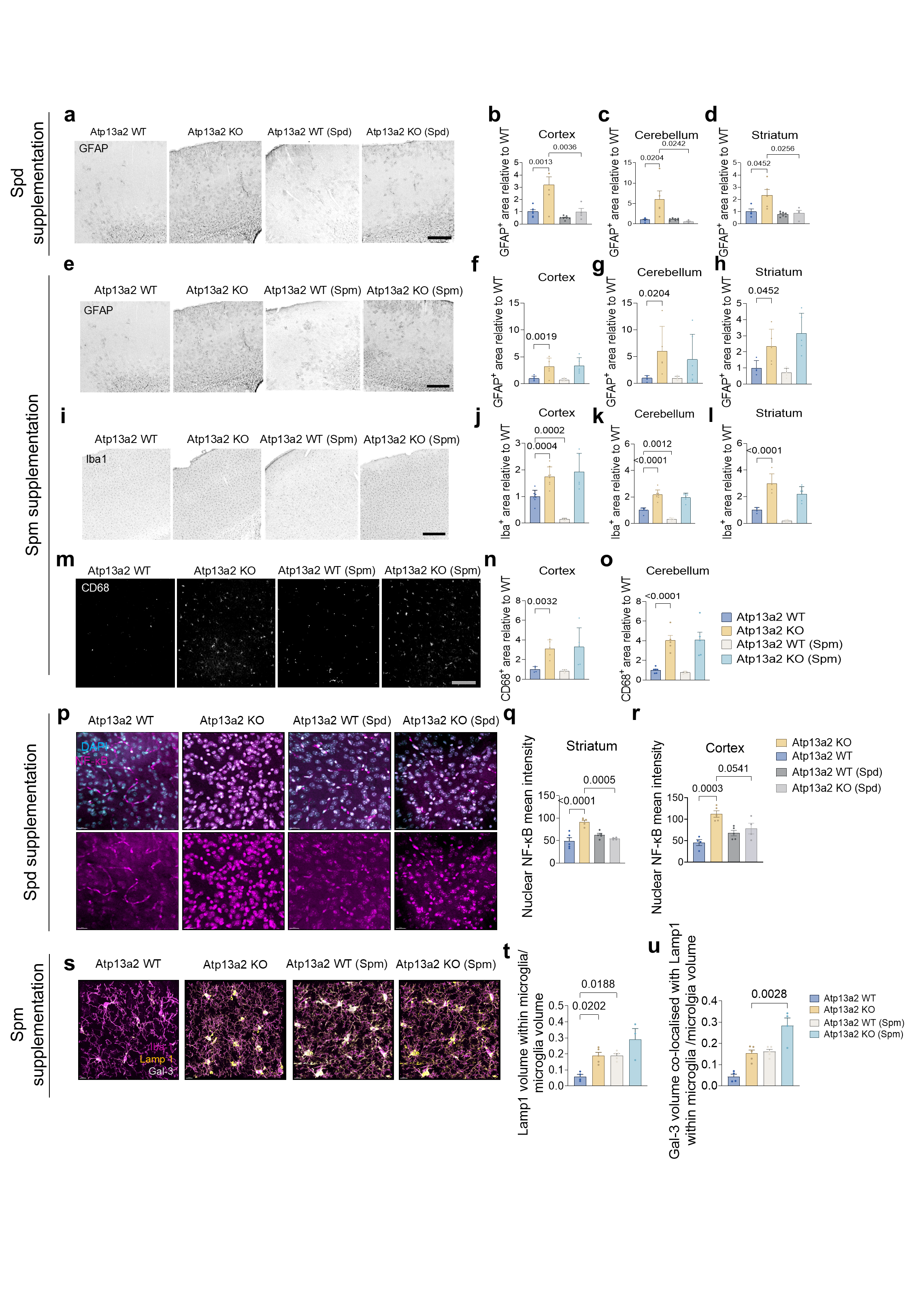

### Supplementary Figure 5

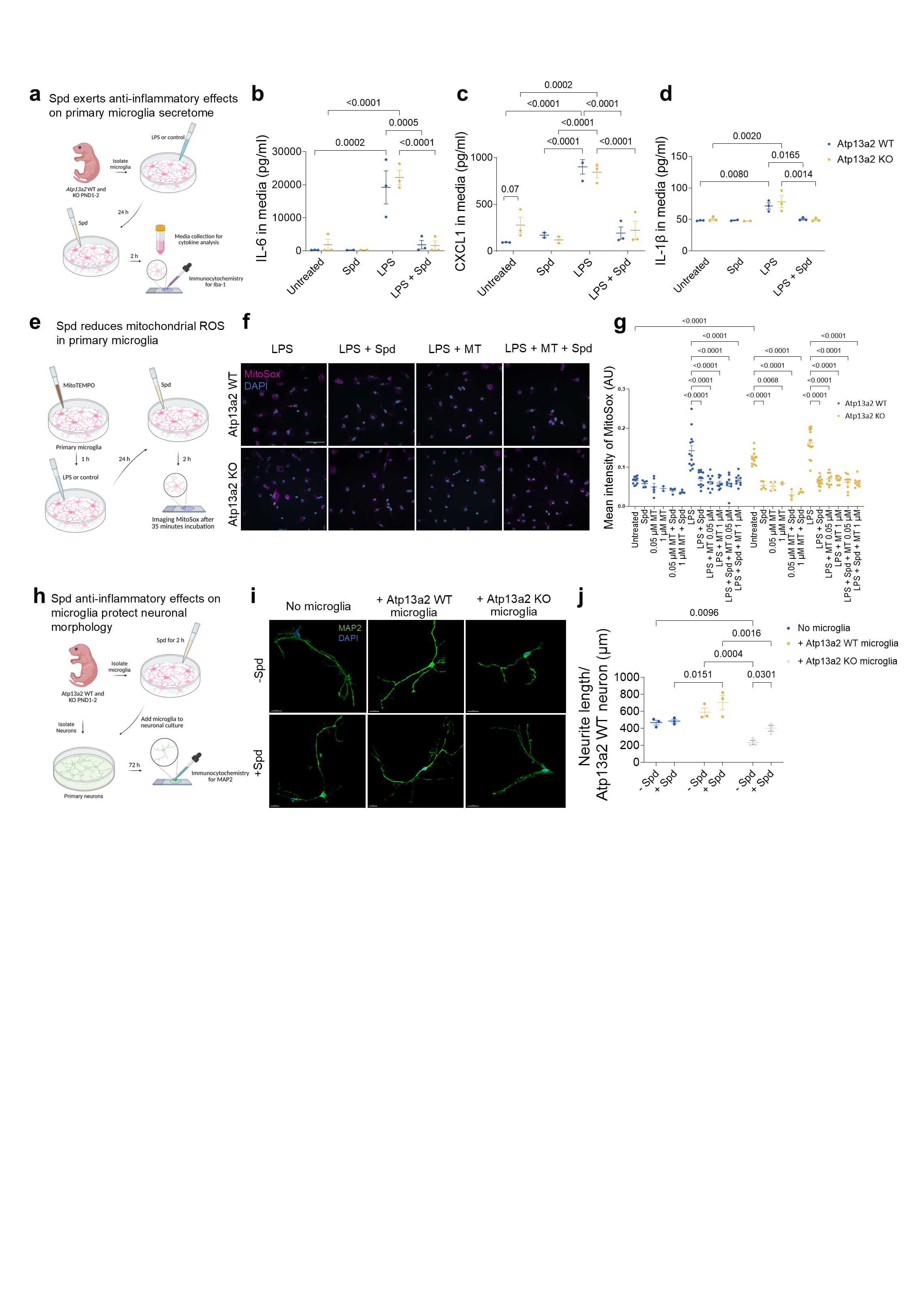
